# Instantaneous Reproduction Number Estimation From Modelled Incidence

**DOI:** 10.64898/2025.12.11.25342071

**Authors:** Robert Challen, Leon Danon

## Abstract

The time-varying reproduction number (*R*_*t*_) is a critical quantity in monitoring an infectious disease outbreak. We propose a new method for estimating *R*_*t*_ from an infectivity profile, expressed as a generation time distribution, and a time series of probabilistic estimates of disease incidence, modelled as log-normally distributed random variables. This is a common output of disease incidence models that are based on Poisson or negative binomial regression of case counts with a logarithmic link function. The method is deterministic, computationally inexpensive and propagates inherent uncertainty in incidence estimates. We validate the method when applied to the output of two simple statistical incidence models, and using simulated data with a defined *R*_*t*_ and infectivity profile. This combination produces comparable outputs to the de-facto standard ‘EpiEstim’. The method can be applied to estimates of disease incidence from a wide variety of incidence models, including those derived from weekly case counts, or that account for right censoring in observed data.

**Author summary:** In our experience estimating the reproduction number during the COVID-19 pandemic, we found that estimating the incidence rate was a useful first step to correct for artefacts and biases in the raw count data, and to estimate the exponential growth rate. With modelled incidence estimates available, and correcting data issues, we wanted to use them to derive the time-varying reproduction number to help monitor the state of the pandemic. We present a mathematical method and supporting software to estimate *R*_*t*_ from modelled incidence estimates, rather than raw count data, and which is readily applicable to many incidence models.

## Introduction

Estimating the time-varying reproduction number (*R*_*t*_) is an important part of monitoring the progression of an epidemic, informing short-term projections of the epidemic size and hence guides decisions on policy interventions targetting behaviour [1]. Changes in *R*_*t*_ can reflect significant events in a pandemic such as the emergence of novel variants [2], and proved highly significant during the COVID-19 pandemic. The confidence of estimates of *R*_*t*_ in an exponentially growing epidemic plays a important role in policy decisions, and appropriate levels of uncertainty are needed. A detailed review of the reproduction number highlights the difference between instantaneous and case reproduction numbers [3]. The case reproduction number is the average number of secondary cases that arise from individuals infected today, and can only be estimated once these secondary cases have occurred. The instantaneous reproduction number estimates the number of primary infections in the past that have resulted in the secondary infections observed today, and hence is used for real time pandemic monitoring [1]. We concentrate solely on the instantaneous reproduction number in this paper. The canonical framework for estimation of the instantaneous reproduction number, based on renewal equations, direct from case data is the Cori method, as implemented in the R package ‘EpiEstim’ [4, 5].

Beyond ‘EpiEstim’, *R*_*t*_ estimation may be done using a range of techniques with different strengths and weaknesses [5–17], the majority of which are based on a time series of count data reflecting the incidence of infection in the population. Such count data may be new infections, hospitalisations or deaths, and are well known to exhibit specific biases due to incomplete ascertainment, reporting delay, and right truncation, along with more generic data quality issues such as missing values, anomalous values [18], or may only be available as time aggregated data. Another key requirement for the estimation of the reproduction number is a profile of the delay between primary and secondary infections. This is described as the infectivity profile, a time dependent probability distribution, and is equivalent to the generation time distribution [1, 19]. The timing of sequential infections measured by the generation time is not directly observed, so the temporal distribution of the serial interval between the positive test results, or symptom onsets, of known infector-infectee pairs is often used as a proxy [5, 19].

In frameworks such as ‘EpiEstim’, *R*_*t*_ estimates are made direct from count data as a proxy for infection incidence, and this makes it difficult to correct for the issues mentioned above, and in some circumstances difficult to produce appropriate confidence intervals. Model based estimates of infection incidence (*I*_*t*_) commonly use count data and a model based around a time varying Poisson rate (*λ*_*t*_), and using a logarithmic link function. In this common situation, the estimate of the Poisson rate at any given time point (*t*) is a log-normally distributed quantity defined by parameters *µ*_*t*_ and *σ*_*t*_, which include a representation of the uncertainty in the count data. It is appealing to use such a modelled incidence estimate as the basis for an estimate of *R*_*t*_, and include this uncertainty into *R*_*t*_ estimates. Incidence models can be derived in a number of ways, that can correct for biases present in the count data, they are easily inspected for error and can be made tolerant of missing values and outliers, or use temporally aggregated data.

This paper presents a mathematical approach to estimating the instantaneous reproduction number from modelled incidence rather than count data, given an estimate of the infectivity profile, which we refer to as ‘*R*_*t*_ from incidence’. This method propagates incidence model uncertainty and infectivity profile uncertainty into estimates of the reproduction number. It decouples incidence modelling and *R*_*t*_ estimation, which allows correction of biases and data quality issues before *R*_*t*_ estimation.

Supporting implementations of all methods described here are provided in the associated R package “ggoutbreak” (https://ai4ci.github.io/ggoutbreak/). We validate the method using a simulation based on a branching process model with fixed infectivity profile and parametrised reproduction number, coupled with two simple incidence models, and compare the output to reproduction number estimates using the Cori method implemented in the R package ‘EpiEstim’ [5] direct from simulated count data.

## Materials and methods

### Mathematical analysis

To use a modelled estimate of incidence to predict *R*_*t*_ we need to propagate uncertainty in incidence into our *R*_*t*_ estimates. To calculate *R*_*t*_ we can use the backwards-looking renewal equations [1] which incorporate the infectivity profile of the disease (*ω*) at a number of days after infection (*τ*):

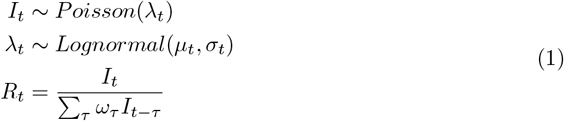

If *k* is the length of the infectivity profile (|*ω*|), in expectation, this gives:

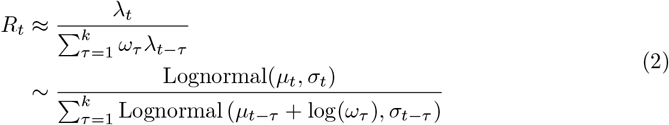

As an aside, it has been shown that the sum of correlated log-normal distributed random variables can be approximated by another log-normal [20] with parameters *µ*_*Z*_ and *σ*_*Z*_, where the correlation between them is *ρ*_*ij*_ = Corr(log(*X*_*i*_), log(*X*_*j*_)). *S*+ in this approximation is the sum of the means of the component log-normals:

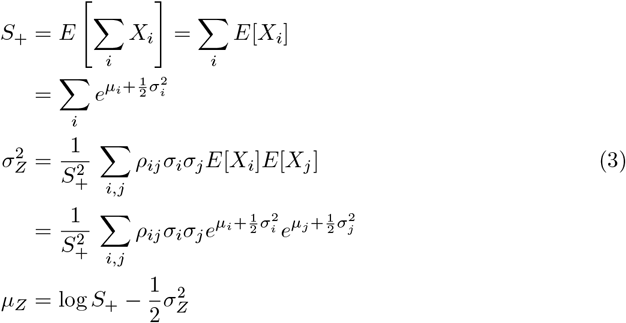

We can apply this approximation (3) to the problem of estimating *R*_*t*_ using the renewal equation. The sum term in the denominator of the renewal equation (2) consists of a set of correlated scaled log normal distributions with scale defined by the infectivity profile (*ω*). For our case for a given time point *t* we equate *X*_*i*_ = *ω*_*τ*_ *λ*_*t*−*τ*_, and substitute *µ*_*i*_ = *µ*_*t*−*τ*_ + *log*(*ω*_*τ*_) and *σ*_*i*_ = *σ*_*t*−*τ*_ into (3) to account for the infectivity profile. We define *k* to be the support of the infectivity profile (*k* = |*ω*|). *m*_*s*_ is the weighted contribution from incidence estimates on day *t* − *τ*, and 𝕍 _*ij*_ is the covariance between log-incidence estimates from days *t* − *i* and *t* − *j*.

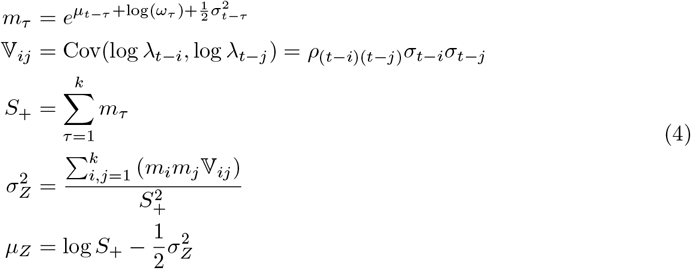

With *µ*_*Z*_ and *σ*_*Z*_ defined, *R*_*t*_ is approximated as the ratio of two log-normals where 𝕍_0*Z*_ = Cov(log(*λ*_*t*_), log(*S*)) is the covariance between the numerator and the log-denominator. Since *S* = ∑_*τ*_ *X*_*τ*_, and using a first-order approximation, this covariance is a weighted average of the covariances between log *λ*_*t*_ and each log *λ*_*t*−*τ*_, weighted by the relative expected contributions *m*_*τ*_ :

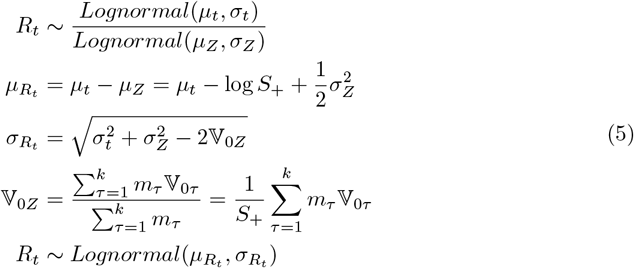

The formulation of *R*_*t*_ in (5) assumes knowledge of the posterior or prediction covariance of the incidence estimates (𝕍_*ij*_). This is typical in modern modelling frameworks [21, 22], but in other situations may not be available. If not available, we could assume the individual estimates of the incidence are independent, however this alters the uncertainty of our *R*_*t*_ estimate and in certain circumstances introduces a potential underestimation bias, influenced by the true off-diagonal mass in the correlation matrix, and the certainty of the incidence estimates. An alternative approach is to assume weak stationarity and estimate a parametric correlation model from the data used to build the incidence model, using Pearson residuals to parametrise an exponential decay function based on time difference [23] (see supporting software package for implementation). The degree of bias involved in the assumption of independence is investigated further in S3 Appendix, and heuristics for assessing the significance of this bias are proposed.

The method for estimating *R*_*t*_ from modelled incidence has been described assuming a non-negative component to the infectivity profile, as it is implicit that infector and infectee are necessarily sequential in time. In the situation where symptomatic case counts are used as a proxy for incidence and the serial interval as a proxy for the infectivity profile, negative times between serial cases may be observed due to variation in delay in observation of the transmission chain. There is nothing in this framework to stop the use of a negative time for the infectivity profile, and we can directly support *R*_*t*_ estimates in these cases.

### Numerical stability

In (5), *µ*_*t*_ is the log-scale mean of the incidence estimate at time *t*, and *σ*_*t*_ its standard deviation. These can be large, leading to numerical instability in terms involving exp(*µ* + *σ*^2^). However, assuming non-negative correlations and using log-space computation with optimized log-sum-exp functions [24], the expressions remain computationally tractable:

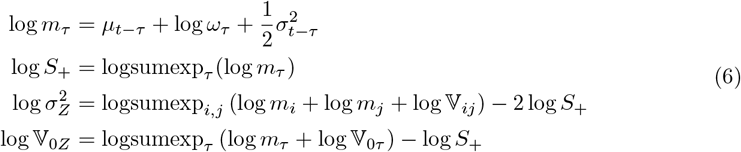

Other relations may be implemented directly as in (5).

### Infectivity profile uncertainty

This estimate of *R*_*t*_ is conditioned on a single known infectivity profile. In reality there is also uncertainty in the infectivity profile (*ω*) which plays a role in the definition of *µ*_*Z,t*_ and *σ*_*Z,t*_. We cannot assume any particular distributional form for the infectivity profile, but we can use a range of empirical estimates of the infectivity profile to calculate multiple distributional estimates for *R*_*t*_ and then combine these as a mixture distribution.

The nature of this mixture distribution will depend on the various estimates of the infectivity profile distributions. However, we can use general properties of mixture distributions to create estimates for the mean and variance of the reproduction number estimate 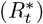 combining the uncertainty arising from multiple infection profile estimates (Ω) and from the incidence estimate model itself:

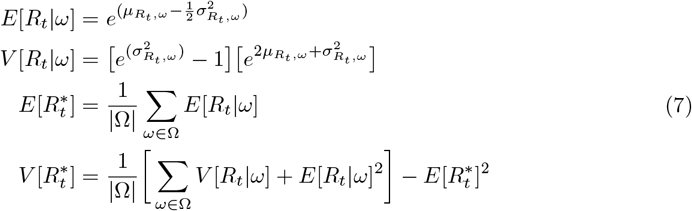

The cumulative distribution function of the mixture is simply the arithmetic mean of the component cumulative distribution functions (conditioned on each infectivity profile). If Φ is the cumulative distribution function of the standard normal distribution:

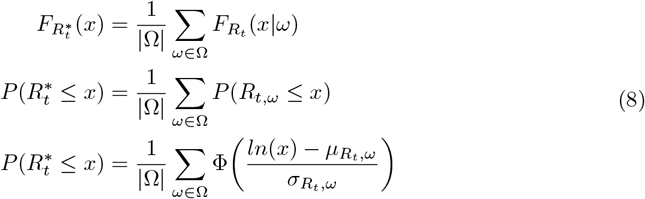

As the cumulative density function of this mixture distribution is a strictly increasing function, specific solutions for median (*q*_0.5_) and 95% confidence intervals (*q*_0.025_ and *q*_0.975_) can be calculated numerically by solving the following equations:

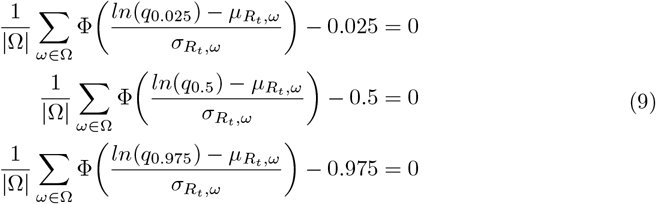

Numerical solutions to this are moderately expensive to perform. A reasonable approximation can be expected by matching moments of a log normal distribution to the mean 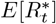 and variance 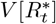 of the mixture. This gives us the final closed form estimator for the reproduction number given a set of infectivity profiles, 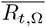, as:

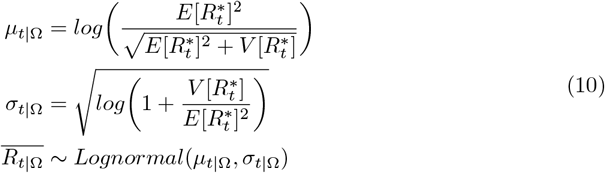

In summary we present a method for retrieving the distributional form of the reproduction number from log normally distributed probabilistic estimates of incidence arising from statistical count models. This includes uncertainty arising from both count models and from infectivity profile distributions.

### Validation

To test this method we developed a simulation based on a branching process model parametrised by 5 different *R*_*t*_ time series, a set number of imported infections at time zero, and fixed infectivity profile (see S1 Appendix Fig S1). Taken together, *R*_*t*_ and the infectivity profile define the expected number of secondary infections given a primary infection, on each day post infection. This expectation is sampled using a Poisson distribution to realise simulated infections on each day. In each simulation run, the degree of outward edges in the network of realised infections at any given time is an instantaneous *R*_*t*_. For each parametrisation of *R*_*t*_ we generate 50 simulations with different random seeds, so that in bulk the simulation reproduction number will be close to the parametrised *R*_*t*_. Each simulation generates a line list of synthetic infections. The line list of infected individuals were aggregated to daily counts of infection. Five random scenarios with different input *R*_*t*_ time series parametrisation were considered, and 50 replicates of each scenario were simulated, with different random seeds, resulting in 250 simulations.

We are particularly interested in uncertainty propagation. To assess the effect of noise in the input case counts in subsequent *R*_*t*_ estimates we assume that infection case counts are subject to varying degrees of ascertainment which change from day to day. The levels of ascertainment were applied to the same underlying infection time series, with observed counts being a binomial sample from the “true” infection counts for any given day. The probability of ascertainment on any given day was a random sample from a Beta distribution with a fixed mean, but three different coefficients of variation (parameter values in the S1 Appendix). In this way there are three versions of each of the 250 simulations which have the same underlying infection counts, but whose case counts only vary by the degree of statistical noise in the observation of infections.

The resulting 750 observed infection counts were used directly as an input to ‘EpiEstim’ to generate a set of baseline *R*_*t*_ estimates, using a window of 14 days. The synthetic infection time-series were also used as input to estimate the underlying infection rate in two ways. Firstly we used a simple statistical Poisson model with time varying rate parameter, represented by a piecewise polynomial of order 2, and fitted using maximum likelihood with a logarithmic link function according to the methods of Loader et al. [25] and a bandwidth equivalent to 14 days as implemented in the R package ‘Locfit’ [26]. We used the central estimate and standard error of this as input to the ‘*R*_*t*_ from incidence’ algorithm, assuming independence. We refer to this estimate as ‘*R*_*t*_ Locfit’. Secondly we estimated the incidence rate using a more sophisticated generalised additive model (GAM) with a smooth time parameter, with regularly spaced knots, one per 14 days, as implemented by the package ‘mgcv’ [23]. We used the central estimates and full covariance matrix of the predictions as input to the ‘*R*_*t*_ from incidence’ algorithm, and we refer to this estimate as ‘*R*_*t*_ GAM’. The resulting sets of estimates of *R*_*t*_ are broadly equivalent to that produced by ‘EpiEstim’, with ‘*R*_*t*_ Locfit’ representing a simple implementation, and ‘*R*_*t*_ GAM’ a more sophisticated implementation of our algorithm.

All three estimators were analysed for estimation delays, by identifying the minimum root mean squared error between estimate and true values when applied to a synthetic dataset designed for this purpose. The lags were corrected for by shifting the *R*_*t*_ estimate by that appropriate number of days (see S1 Appendix Fig S2 and Table S1 for details).

For each estimator method and within the 3 groups of low, medium and high ascertainment noise, 5 scenarios were run with 50 replicates of each scenario. The posterior distributions of daily *R*_*t*_ estimates from both estimators, ‘EpiEstim’, ‘*R*_*t*_ Locfit’ and ‘*R*_*t*_ GAM’, were compared to simulation ground truth at all time points and summarised for each time series to give estimator performance metrics for each of the 750 simulation replicas. From the each of these replicas 20 bootstraps were resampled during summarisation (resulting in 15,000 sets of estimator metrics per method). Estimator metrics for each method were graphically summarised to the 3 groups and presented as box-plots (main paper) and summarised to each of 5 scenarios within each of the 3 groups (full details in S2 Appendix). Comparisons between methods were made graphically.

We calculated the average continuous ranked probability score (CRPS) as a overall performance metric [27–31]. The average proportional bias of the estimates within each simulation gives us a measure of estimator bias. We calculated the mean of the 50% interval width (inter-quartile range) of each estimate as a measure of estimator sharpness. We calculated the coverage probability of the 50% inter quantile range as an indicator of estimator calibration. We further investigated calibration by de-biasing estimates and with these adjusted estimates derived a novel calibration metric as the Wasserstein distance [32] of the probability integral transform histogram from the uniform [33–36] (lower values are better). To test the ability to discriminate between a growing or shrinking epidemic we calculated the prediction probability of misclassification of the true value *R*_*t*_ being greater than or less than 1. A weighted average of these by absolute distance of the true value from 1 gives us a estimator metric for this specific question (lower values are better). These metrics are fully defined in S2 Appendix.

We conducted two sensitivity analyses, one with an ‘EpiEstim’ window and ‘Locfit’ bandwidth. or ‘GAM’ knots equivalent to 7 days, rather than the 14 in the main analysis, and a second comparing estimate quality when the estimates were not corrected for delays.

## Results

In Fig 1 panel A case counts and a modelled incidence estimate from a single simulation are shown for the 3 levels of ascertainment noise. Uncertainty in modelled incidence estimates increases with noise. In panel B we compare the *R*_*t*_ estimates derived from this this simulation. ‘EpiEstim’ can be observed to produce a slightly lagged estimate (top row panel B). In general the confidence of ‘EpiEstim’ estimates appear related to the distance of the estimate from 1. The central estimate becomes more volatile with more noise in the data set, but the confidence intervals do not appear to widen, suggesting noise in the input data does not affect uncertainty. By comparison ‘*R*_*t*_ GAM’ estimates (second row panel B) are smoother, less lagged and suffer from fewer tail effects at the limits of the time series. ‘*R*_*t*_ Locfit’ estimates (lower row panel B) also show no obvious lag, are overall more uncertain, particularly at the start and end of the time series. Increased noise in the data increases the uncertainty in the ‘*R*_*t*_ Locfit’ estimates more than the other two estimates.

**Fig 1.**
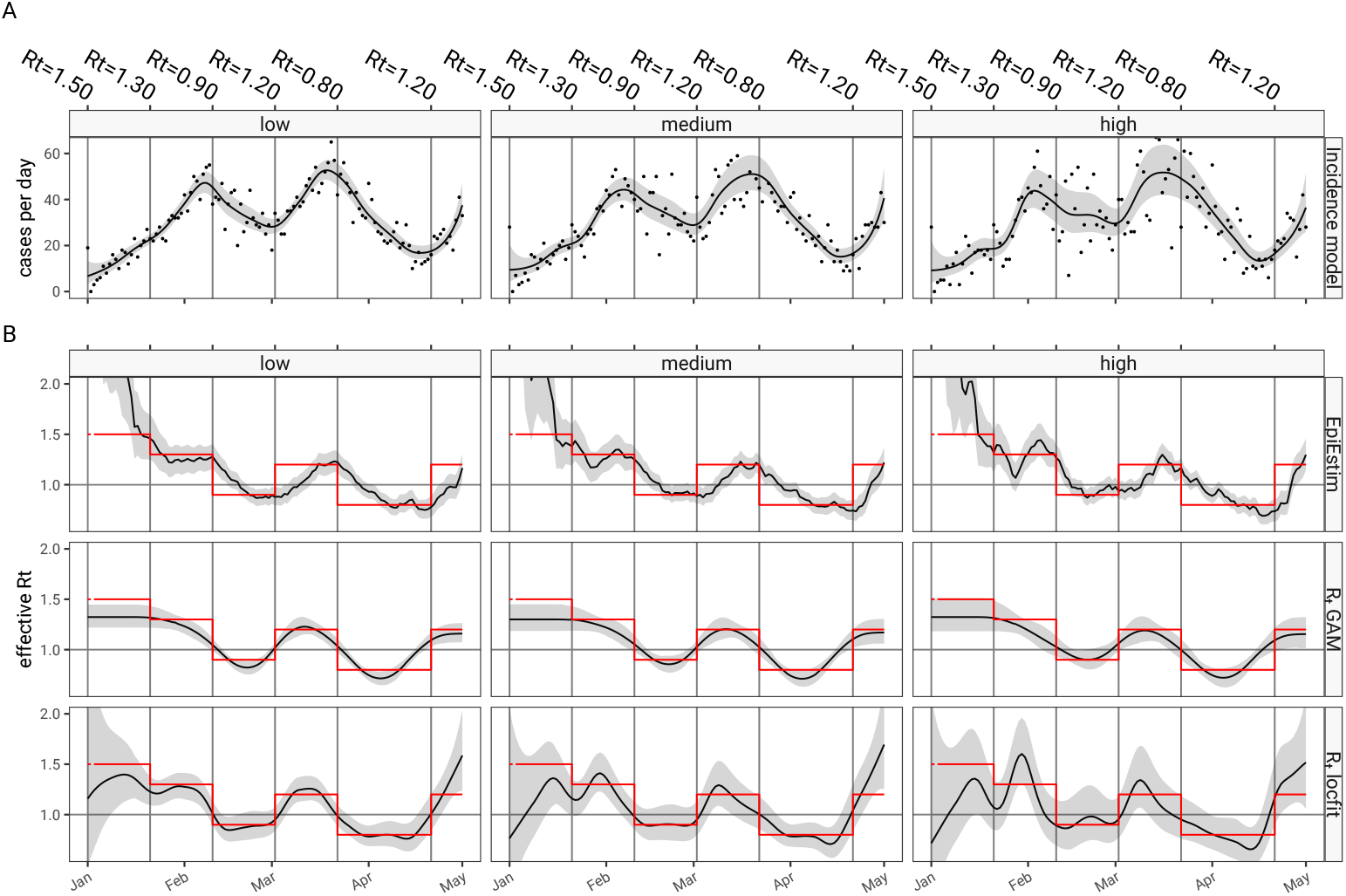
Instantaneous reproduction number estimates from a branching process model simulation. A qualitative comparison of instantaneous reproduction number estimates is shown. Panel A shows three case time series based on a single run of a branching process model parametrised with a stepped reproduction number time series (red lines in panel B) and infectivity profile as in S1 Appendix Fig S1. Case counts are shown as dots. A smoothed estimate of the cases per day as a line with shaded 95% confidence intervals, based on a simple Poisson regression model. All three time series have on average 70% case ascertainment, however the day to day variability of ascertainment is parametrised as a Beta distributed random variable, with “low”, “medium” and “high” relating to the coefficient of variation of the Beta distribution (see S1 Appendix Table S1). Panel B shows estimates of the reproduction number based on the methods presented in this paper, and in the top row ‘EpiEstim’ estimates derived from the data points in panel A are shown. In the middle row *R*_*t*_ esimtates from a combination of GAM incidence model and the methods described in the paper. In the bottom row, *R*_*t*_ estimates derived from a ‘Locfit’ incidence model. In panel B the parametrised *R*_*t*_ is shown as a solid red line and can be regarded as the ground truth for this single simulation run.

In the main validation scenario we used a window for ‘EpiEstim’, ‘GAM’ and ‘Locfit’ of 14 days. We saw in Fig 1 that this results in the estimate of *R*_*t*_ lagging the true value. This is quantified in Table 1 and Fig S2 in the S1 Appendix. In the main analysis ‘EpiEstim’ tends to produce an estimate delayed by 7 days and this lag is corrected by shifting the estimate in time before other metrics are calculated. In the first sensitivity analyses with a window of 7 days, a 4 day lag is observed for ‘EpiEstim’, and in the second sensitivity analysis the metrics are calculated without correcting for lags.

**Table 1.**
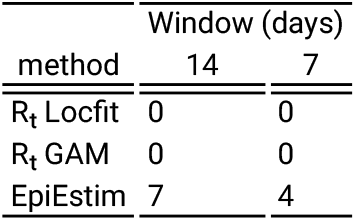
Estimator delays in the validation scenarios.

| method | Window (days) |  |
| --- | --- | --- |
|  | 14 | 7 |
| $R_t$ Locfit | 0 | 0 |
| $R_t$ GAM | 0 | 0 |
| EpiEstim | 7 | 4 |

A quantification of the quality of the two estimation methods is shown in Fig 2 summarised from all 750 simulations, and corrected for estimator delays. In panel A the continuous ranked probability score is lower (better) for ‘*R*_*t*_ GAM’ and ‘*R*_*t*_ Locfit’ over all scenarios combined. In panel B the proportional difference between the true value and the median of the estimate probability (expressed as a percentage change) shows that the ‘EpiEstim’ and ‘*R*_*t*_ GAM’ exhibit little bias, whereas ‘*R*_*t*_ Locfit’ demonstrates a tendency to underestimation, in this set of experiments. In Fig 2 panel C, the 50% prediction interval width is smaller for ‘EpiEstim’ and ‘*R*_*t*_ GAM’ demonstrating sharper, more confident, estimates than the predictions of ‘*R*_*t*_ Locfit’. ‘*R*_*t*_ Locfit’ is the only method which has evidence of decreasing sharpness (increasing interval width - Fig 2 panel C) with increasing data noise. ‘EpiEstim’ does not exhibit this change in sharpness with data noise and ‘*R*_*t*_ GAM’ only minimally. The low values of the probability of 50% prediction coverage, as seen in panel D, which is ideally 0.5, implies that ‘EpiEstim’ and ‘*R*_*t*_ GAM’ are over-confident in its predictions, ‘*R*_*t*_ Locfit’ on the other hand looks to be somewhat conservative, at least with more noisy data. The PIT Wasserstein metric in Fig 2 panel E, allows us to compare the calibration of the 2 methods on the same scale, one of which is over confident and the other being under confident. This suggests ‘*R*_*t*_ Locfit’ is better calibrated than ‘EpiEstim’ or ‘*R*_*t*_ GAM’ (albeit in a different direction). Finally Fig 2 panel F shows the probability of misclassification at the threshold of *R*_*t*_ = 1 which is similar for ‘EpiEstim’ and ‘*R*_*t*_ Locfit’, but lower (better) for ‘*R*_*t*_ GAM’. We also looked at how these metrics varied between scenarios (details in S1 Appendix Fig S3 and S4) and the breakdown largely follows the summary presented in Fig 2, although ‘*R*_*t*_ GAM’ performs relatively poorly in scenario 3, and in scenario 5, which has relatively little variation compared to the others, calibration is better for ‘EpiEstim’, and ‘*R*_*t*_ GAM’ appears overconfident.

**Fig 2.**
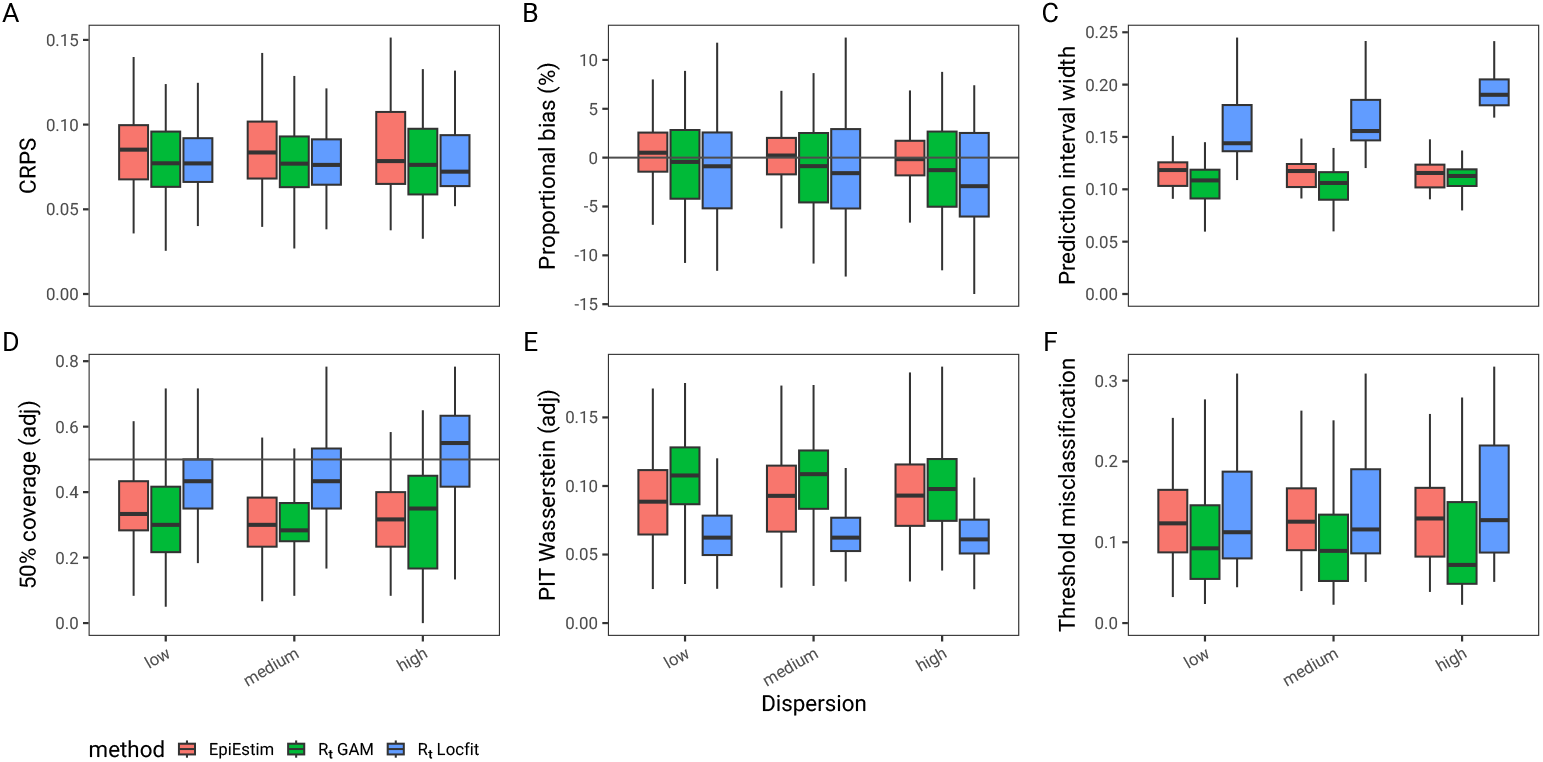
Quantitative comparison of *R*_*t*_ estimation methods applied to 50 simulations of 5 scenarios at 3 levels of ascertainment noise. The figure compares metrics describing the overall performance of the estimators. In Panel A is the continuous ranked probability score (CRPS) - lower is better; the average proportional bias (panel B) which characterise bias - lower is better; In panel C, the 50% prediction interval width measures estimator sharpness, and lower is better if the estimator is unbiased and well calibrated. In Panel D the probability of 50% coverage (ideal value is 0.5), and in Panel E the adjusted probability integral transform (PIT) Wasserstein metric (lower is better) are both measures of calibration. In Panel F a functional metric which quantifies the probability that an estimate is the wrong side of the cirtical threshold of *R*_*t*_ = 1 desribes utility in decision making.

In the first sensitivity analysis (S1 Appendix Fig S5), the amount of data informing the *R*_*t*_ estimates is reduced from 14 to 7 days, resulting in reduced certainty in both estimators. This changes the relative performance of the 2 methods as measured the the CRPS, which is now tends to favour ‘*R*_*t*_ GAM’ as the better overall estimator, despite continued bias, excess sharpness and mis-calibration. The certainty of ‘*R*_*t*_ Locfit’ has dropped to a low level and it has become excessively conservative in all but the high ascertainment noise scenarios (further details in S1 Appendix). This also slightly increases the probability of misclassification at the threshold of *R*_*t*_ = 1 for ‘*R*_*t*_ Locfit’, although ‘*R*_*t*_ GAM’ still outperforms other methods on this more functional metric. In the second sensitivity analysis with no correction for lag (details in S1 Appendix Fig S6), ‘EpiEstim’, which is affected by lag, performs much less well in all metrics as the lag penalises all dimensions of the quality of the estimator, and this serves to highlight the importance of addressing lag before comparing the bias and calibration of estimators.

## Discussion

In this paper we describe a mathematical method for deriving an estimate of the effective reproduction number (*R*_*t*_) from modelled estimates of disease incidence, that incorporates uncertainty from both incidence model and infectivity profile. By adopting a two stage process it allows for a flexible approach to incidence modelling that allows us to address many issues, such as right truncation, anomalies, missing data, or temporally aggregated data, before using this to estimating *R*_*t*_. At the same time our method propagates uncertainty from incidence models and infectivity profiles appropriately. We provide evidence that when combined with basic incidence models, our method produces *R*_*t*_ estimates that are comparable to the de-facto standard algorithm implemented in ‘EpiEstim’ [5], when assessed against synthetic outbreak data.

There is a similar approach to deriving *R*_*t*_ from modelled incidence described by Gressani et al. [13]. It assumes a specific formulation for the incidence model as a negative binomial distribution estimated with P-splines in a Bayesian framework with Laplace approximations. This encouragingly also gives log normally distributed estimates for *R*_*t*_, but its methodology is predicated on the specifics of the P-spline posterior vector and it does not incorporate infectivity profile uncertainty.

By comparison the approach in this paper is only loosely coupled to the incidence estimate framework and so can be applied to any incidence model that produces a time varying log-normally distributed incidence estimate. This includes a broad family of Poisson and negative binomial regression models [21, 25, 37], or latent Gaussian models [22] using logarithmic link functions, and is agnostic to the formulation of those models. The incidence models can be estimated on time aggregated data [11], include covariates such as day of week effects, incorporate change points without affecting the derivation of the reproduction number estimates.

In terms of infectivity profile, our method is robust to distributions with zero or negative time intervals between index and secondary observations. It could therefore be used with directly observed real world serial interval distributions [19] and delayed case counts as proxies for infectivity profile and infection incidence, which are necessarily inferred quantities; this is explored further in the ‘ggoutbreak’ package documentation (https://ai4ci.github.io/ggoutbreak/). Our method does not specifically address important questions arising from ascertainment bias, or right truncation of observed incidence [6, 18], however when input incidence models have been adapted to right censored data our method can be used to derive an *R*_*t*_ estimate (see package documentation for more details).

The validation comparison here combines one very simple statistical model of incidence ‘*R*_*t*_ Locfit’, and one more sophisticated model ‘*R*_*t*_ GAM’, with our method to derive *R*_*t*_ estimates, and compares them to estimates produced direct from the count data by ‘EpiEstim’. This shows that the combination of incidence models and our *R*_*t*_ derivation, produces estimates similar to ‘EpiEstim’. We have not formally tested the statistical significance of the differences because they are based on a large number of observations, and even tiny differences will be statistically significant. We pragmatically chose to simulate using a set of 5 step functions for *R*_*t*_ parametrisation, which we expect to be relatively challenging for both ‘EpiEstim’ and the statistical models for incidence we used here. It is clear that relative performance between the methods varies with the exact details of the test scenario (see S1 Appendix Fig S6). A more realistic smooth *R*_*t*_ parametrisation time series has been performed and both methods perform better in such scenarios, so we consider our simulations to be worst case. There are some scenarios in which ‘EpiEstim’ performs better and others in which incidence model derived estimates perform better. In comparing the models we ignored the first 20 *R*_*t*_ estimates which are unstable in ‘EpiEstim’ and very uncertain in ‘*R*_*t*_ Locfit’, this will tend to discriminate against ‘*R*_*t*_ GAM’ which seems comparatively accurate in the first 20 days.

Our method is not a replacement for ‘EpiEstim’ as it requires an incidence model dervied from count data, rather than directly using data, and any comparison is looking at both the incidence model quality and the method for deriving *R*_*t*_. This must be taken into account when interpreting the validation section of this paper. By picking two statistical models for incidence with different characteristics, to which to apply our method for estimating *R*_*t*_, we hope to demonstrate that the combinations are not obviously inferior to ‘EpiEstim’. If we had used different incidence models, the overall *R*_*t*_ estimate may have very different characteristics. The choice of best estimator is subjective as it depends on whether it is more important to have an estimator that minimises the risk of a misclassification between a growing or shrinking epidemic, or whether a more accurate representation of uncertainty is required. Estimates of later time points are also arguably more important in managing an epidemic.

Notwithstanding these limitations in validation, we argue our method for deriving *R*_*t*_ from log-normally modelled incidence estimates, which are commonly produced by statistical modelling frameworks, is a useful adjunct to the range of tools available for monitoring an epidemic. It is relatively quick and deterministic, and is flexible enough to be combined with a wide range of temporal incidence modelling techniques, which can account for reporting delays or ascertainment bias, or could be extended to spatio-temporal incidence models.

## Funding

RC and LD are funded by UK Research and Innovation AI programme of the Engineering and Physical Sciences Research Council, AI for Collective Intelligence Research Hub (EPSRC grant EP/Y028392/1; https://gtr.ukri.org/projects?ref=EP%2FY028392%2F1). RC and LD are affiliated with the JUNIPER partnership funded by the Medical Research Council (MRC grant MR/X018598/1; https://www.ukri.org/councils/mrc/). The views expressed are those of the authors.

## Competing interests

The authors have no competing interests to declare.

## Author contributions

RC and LD generated the research questions. RC performed the mathematical analysis and simulations, and created the supporting software package. RC and LD provided validation of the methods. LD provided supervision of the research. RC developed the first draft of the manuscript. RC and LD contributed to the final editing of the manuscript and its revision for publication and had responsibility for the decision to publish.

### Use of large language models

We acknowledge the input of the large language model QWEN3-235B-A22B-2507, principally in refining the mathematical methods in this paper. All methods were conceptualised by the authors, but QWEN3 contributed to improve the handling of estimate covariance and assessment of the bias when assuming independence (S3 Appendix). QWEN3 was also used to improve mathematical notation and consistency. There was no use of LLMs in generating the final narrative of this paper, or in validation of the methods. The reference implementations of these methods was written without the use of LLMs, except for the code responsible for the generation of synthetic variance-covariance matrices. All mathematical and code generated by QWEN3 was rigorously reviewed and tested by the authors. Discussion history is available from the corresponding author on request.

## Data and code availability

All data and code used for running experiments, model fitting, and plotting is available on a GitHub repository at https://ai4ci.github.io/ggoutbreak-paper/. The methods described here are implemented in the form of an R package to support the estimation of epidemiological parameters and it is deployed on the AI4CI r-universe (https://ai4ci.r-universe.dev/ggoutbreak). We have also used Zenodo to assign a DOI to the repository: doi:10.5281/zenodo.7691196.

## Supporting information

**S1 Appendix. Simulation for validating Rt estimates, additional results and sensitivity analyses** Methodological details of simulation set up for validating the performance of *R*_*t*_ estimators presented in the main paper, additional figures and detailed results from the sensitivity analyses.

**S2 Appendix. Metrics for evaluating the quality of probabilistic estimators**. Methodological details of performance metrics used for validating the performance of *R*_*t*_ estimators presented in the main paper.

**S3 Appendix. Bias in** *R*_*t*_ **Estimation Under Independence and Heuristics for Risk Assessment**. This appendix provides bounding of bias in *R*_*t*_ estimates under the assumption of independence of incidence estimates, and derives of metrics to assess whether the assumption of independence is valid.

## Notes

### Competing Interest Statement

The authors have declared no competing interest.

