## Supplementary material for "Instantaneous Reproduction Number Estimation From Modelled Incidence": S1 Appendix

### Supplementary 1: Simulation for validating $R_t$ estimates, additional results and sensitivity analyses

#### 1 Introduction

In the main paper we develop an algorithm for the estimation of the reproduction number $R_t$, and show that it produces sensible estimates for $R_t$ by using an infectious disease outbreak simulation parametrised with $R_t$. This supplementary provides detail the on the approach to simulation and validation of the $R_t$ method described in the main paper, and provides additional figures and the results of sensitivity analyses. Further detail on the performance metrics reported is available in Supplementary Appendix 2.

#### 2 Simulation

The basis for the simulation and subsequent estimation of $R_t$ relies on an infectivity profile, that defines the delay distribution of secondary cases given primary infections [1, 2]. In a real world situation this delay distribution may be inferred from data and associated with uncertainty. To replicate this we generate a set of infectivity profiles based on a gamma distribution with uncertain parameters as described in figure 1. The average value of this distribution is used as the "true" value and used to generate the simulated data, combined with an $R_t$ parameter. The multiple possible infectivity profiles in Fig 1 represent the uncertainty we would expect in a real scenario, when infectivity profile is inferred from data. Therefore all 100 infectivity profiles are used in estimating the reproduction number, and contribute to its uncertainty, however the same central infectivity profile is used during all simulation runs.

The simulation is a stochastic discrete time branching process model where each node in the model is an infected individual. The model is parameterised by a time varying reproduction number ($R_t$), and an infectivity profile ($\omega$). The model is seeded with an initial set of 30 infected individuals. Growth of the model is not constrained to a population size. The probability that any infected individual ($X_{i,t}$) who was infected at time $t$ generates secondary infections ($X_{j,i,t+\tau}$) on a subsequent day $t+\tau$ is determined by the reproduction number on that day ($R_{t+\tau}$) and the infectivity profile of the day since primary infection ($\omega_\tau$).

$$E(X_{j,i,t}|X_{i,t},\tau) = \omega_\tau \times R_{t+\tau} \tag{1}$$

From the branching process model the count of simulated infections for each day ($I_t$) is the "true count" of infections. For the purposes of the simulation this true count is said to be partly observed with a different probability of ascertainment each day ($P_{asc}$). This probability is a random sample from a Beta distributed quantity with mean of 0.7 and different degrees of

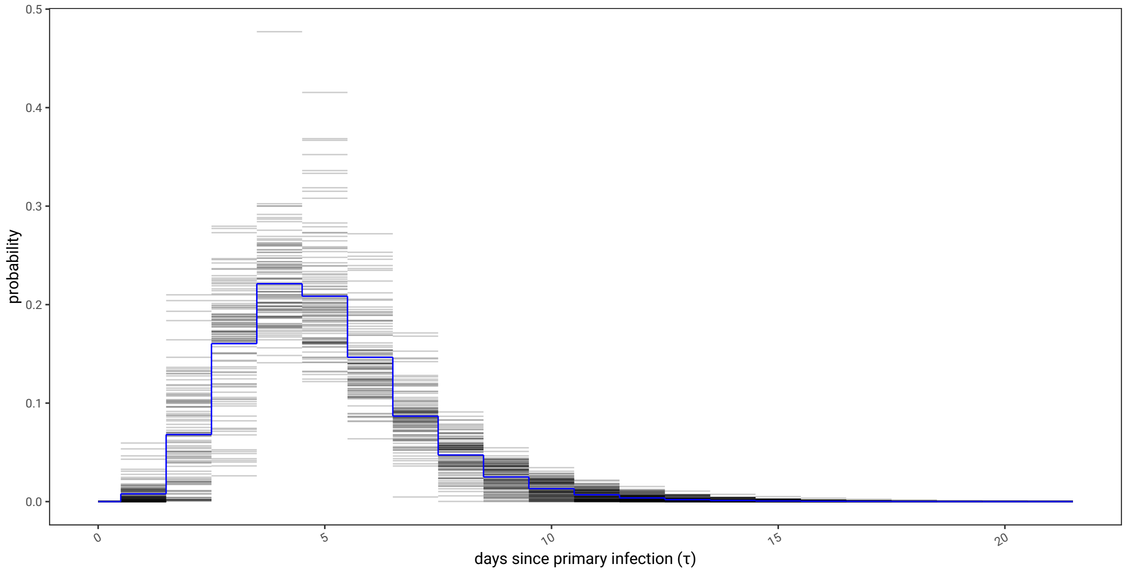


Figure 1: A simulated set of 100 infectivity profiles generated by the R package 'EpiEstim', using mean of 5 (± 1) days and SD of 3 (± 1) days, as an offset gamma distribution, discretised on daily intervals, and truncated at 14 days. The average of these infectivity profiles is regarded as the "true" value and is highlighted in blue.

Table 1: Parameters of a Beta distribution ($\alpha$ and $\beta$) which decide the day to day ascertainment rate of the simulations ($P_{asc}$)

| dispersion | $\alpha$ | $\beta$ | mean: $\mu$ | coefficient of variation: $\frac{\sigma}{\mu}$ |
|---|---|---|---|---|
| low | 302.63 | 129.7 | 0.7 | $3.145 \times 10^{-02}$ |
| medium | 18.25 | 7.83 | 0.7 | $1.258 \times 10^{-01}$ |
| high | 5.49 | 2.35 | 0.7 | $2.201 \times 10^{-01}$ |

dispersion as outlined in the table below (expressed as coefficients of variation), the result of which is an observed count of infections $O_t$:

$$\begin{aligned} I_t &= |X_t| \\ O_t &\sim Binom(I_t, P_{asc}) \\ P_{asc} &\sim Beta(\alpha, \beta) \end{aligned} \tag{2}$$

This observed case count is a fraction of the same infection counts for each scenario replication and differs only by the day to day variation in ascertainment.

### 3 Validation

We compare different methods for estimating $R_t$, 'EpiEstim' and '$R_t$ from incidence' as described in the main paper. The different $R_t$ estimators are subject to varying delays as seen in Fig 1 in the main paper and Fig 2 in this appendix. The lags can be quantified by estimating the reproduction number for a well characterised dataset generated with a periodically varying reproduction number. The minimum root mean squared error between prediction and known $R_t$ is taken as the estimate lag. We see the lag introduced by 'EpiEstim' is half of the window size [1, 3] using the standard interpretation of 'EpiEstim' as the $R_t$ estimate at being at the end of the smoothing window.

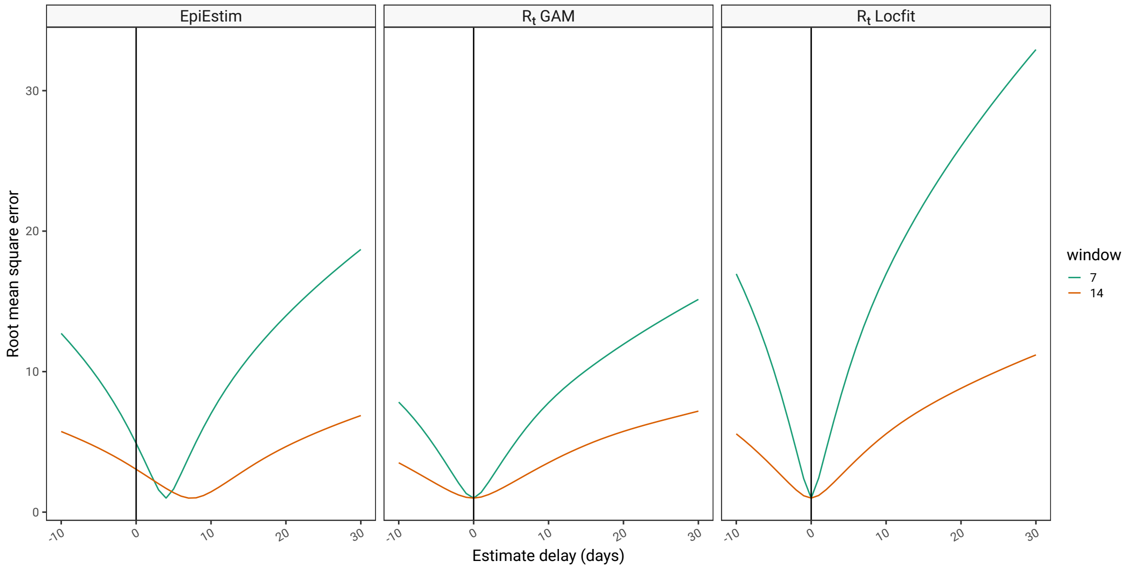


Figure 2: Lags in $R_t$ estimation methods. 'EpiEstim' integrates information over a window, during which $R_t$ is assumed constant. By convention the time point for the estimate is taken to be the end of the window and therefore the estimate is lagged (left panel). The $R_t$ from modelled incidence method described here is only affected by delays in the incidence estimation but the method we chose is not lagged (right panel).

The testing consisted of 50 replicates of 5 randomly generated scenarios, each with 3 levels of ascertainment dispersion. The various scenarios and the highest and lowest $R_t$ estimates for the scenario replications are shown in Fig 3. During the very early phase of an epidemic $R_t$ estimators results are typically unreliable. When comparing estimators we only use the estimates from day 20 to 80, when they have established a pattern. The scenarios (which were randomly generated) are quite similar apart from simulation 5 which has smaller variation.

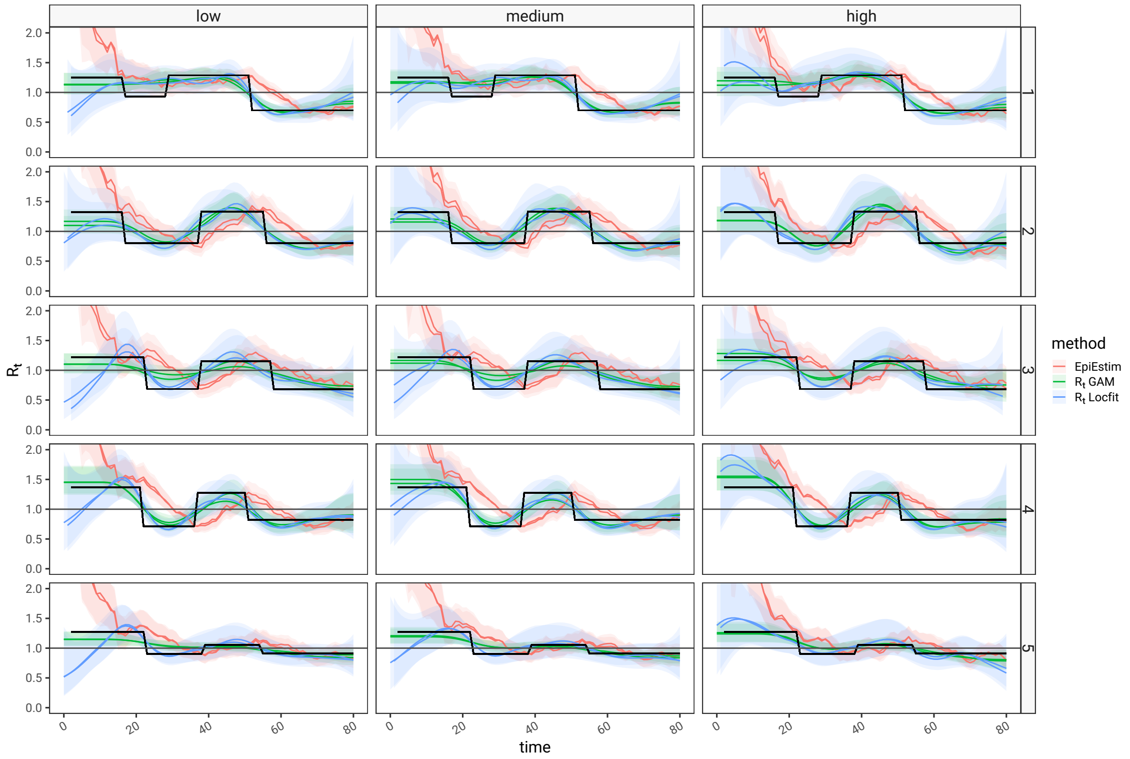


Figure 3: The five scenarios tested. Each scenario (rows) had 50 replications and 3 levels of ascertainment with different dispersion applied to them (columns). The coloured traces show the replicate resulting in the highest and lowest $R_t$ estimates for each scenario, for each method. During validation the first 20 days worth of estimates were discarded as the lead in to the time series predictions suffers from a specific inaccuracy due to the incomplete nature of the data informing the renewal equation in this early phase.

### 4 Additional results

The selection of metrics for validation are introduced in the main paper, and described in detail in Supplementary appendix 2. In this supplementary we show results faceted by scenario and ascertainment bias. These additional results give us some sense as to whether there are specific features of the testing scenario that favour one estimator over the other.

Fig 4 estimator quality stratified by simulation scenario and ascertainment noise shows similar patterns to the main paper. In panel A the mean CRPS shows that in all scenarios the '$R_t$ Locfit' estimator scores lower (better) and in panel B the proportional bias is usually closer to zero (less biased). 'EpiEstim' and '$R_t$ GAM' are consistently sharper than '$R_t$ Locfit' meaning that it produces results with narrower confidence intervals. In the presence of bias this is not always desirable. Calibration is assessed in panel D and '$R_t$ from incidence' estimates are closer to the ideal value of 0.5 in most scenarios, except for scenario 5, this suggests that in scenarios 1-4 'EpiEstim' and '$R_t$ GAM' are over confident, whereas in scenario 5 '$R_t$ Locfit' is excessively cautious. Similarly the other measure of calibration, the adjusted PIT-Wasserstein score (details in supplementary 2) is better (lower) in '$R_t$ Locfit' in scenarios 1-4.

Scenario 5 was noted above to have less variation in high and low values. This suggests that 'EpiEstim' and '$R_t$ GAM' are more accurate when the time series is more stable. Step changes should be equally challenging for both methods to adapt to (which is why they were chosen), and have relevance to behvioural interventions such as lockdown, but other validation scenarios such as smoothly changing $R_t$ values could be considered.

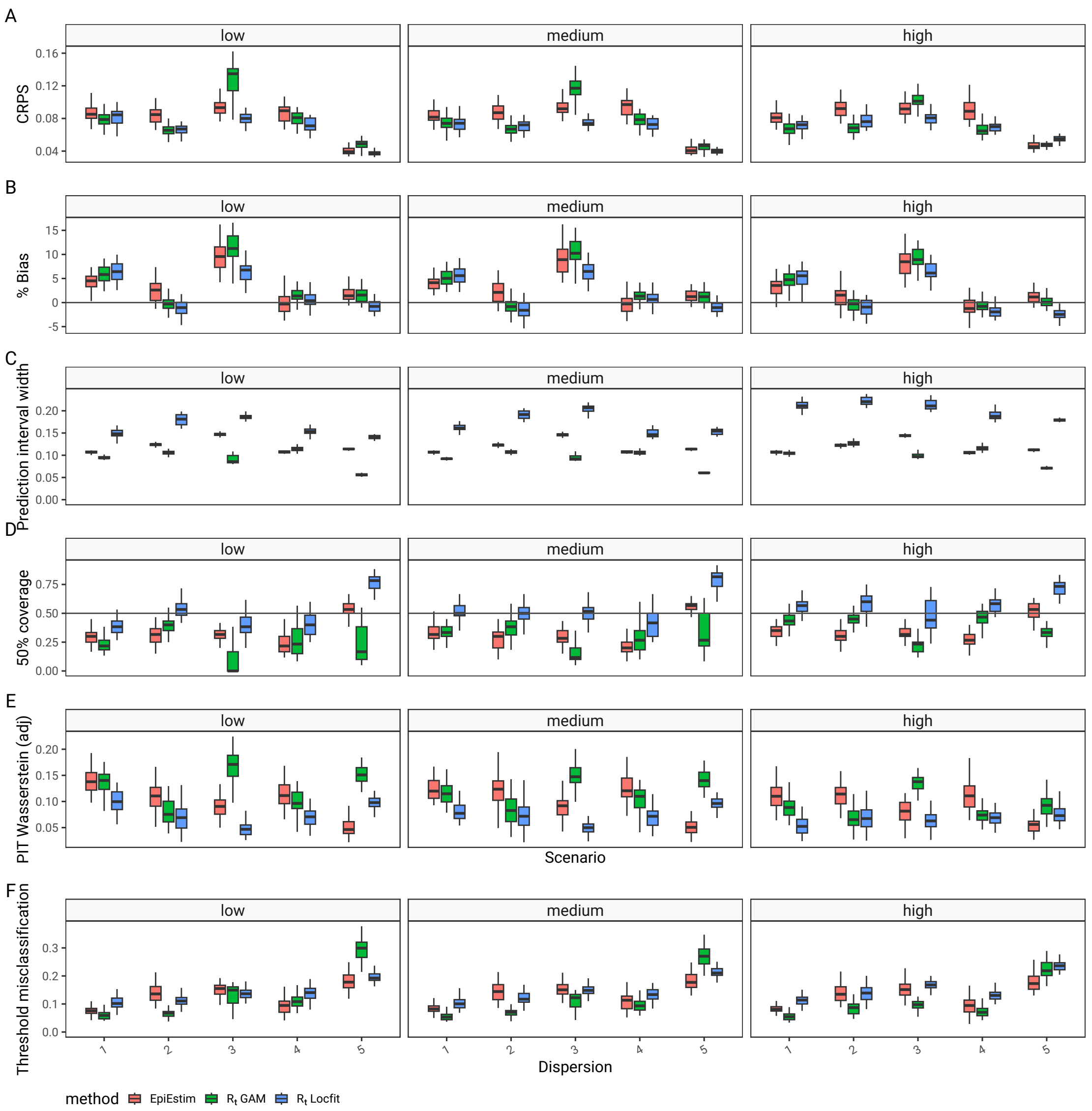


Figure 4: Mean Continuous Ranked Probability Scores (panel A), proportional bias as a percent (panel B), 50% prediction interval width (panel C), 50% coverage probability (panel D) and Probability integral transform (PIT) histogram Wasserstein distance from uniform (Panel E), stratified by scenario and ascertainment noise. Lower values are better in all metrics apart from the 50% coverage probability (panel D) which is ideally 0.5. Lower values of the prediction interval width are only unequivocally better in an unbiased and well calibrated estimator, otherwise a trade off between sharpness, bias and calibration is required.

### 5 Sensitivity analyses

In the first sensitivity analysis we re-ran the 2 estimators with different configuration. In this case we narrowed the data window on which the estimates are based. This made all estimators less certain and more reactive to changes in the data (less smooth). These effects are seen in the prediction time series in Fig 5 panel A. In terms of the quality metrics found in in Panels B-G, the the estimators show a similar behaviour to the main analysis in terms of bias and sharpness (Panel C and D), but 'EpiEstim' is now slightly better calibrated with the increased unceratinty due to the shorter data window (panel E). The overall performance as measured by CRPS is best for '$R_t$ GAM' measured by the CRPS (panel B). The calibration suggests that '$R_t$ Locfit' is now tending towards being excessively conservative, except when ascertainment noise is particularly high (Panel E), whereas other estimates remain excessively certain. In terms of overall misclassification at the threshold of $R_t = 1$ there is still not much to choose between the 'EpiEstim' and '$R_t$ Locfit'm, but '$R_t$ GAM' is better than both.

In the second sensitivity analysis (see Fig 6) we repeat the analysis on the main paper but without adjusting for estimator lag before calculating the metrics. This does not change the findings in the main paper, as if anything it penalises 'EpiEstim' unfairly due to fact that the '$R_t$ from incidence' does not suffer from lag.

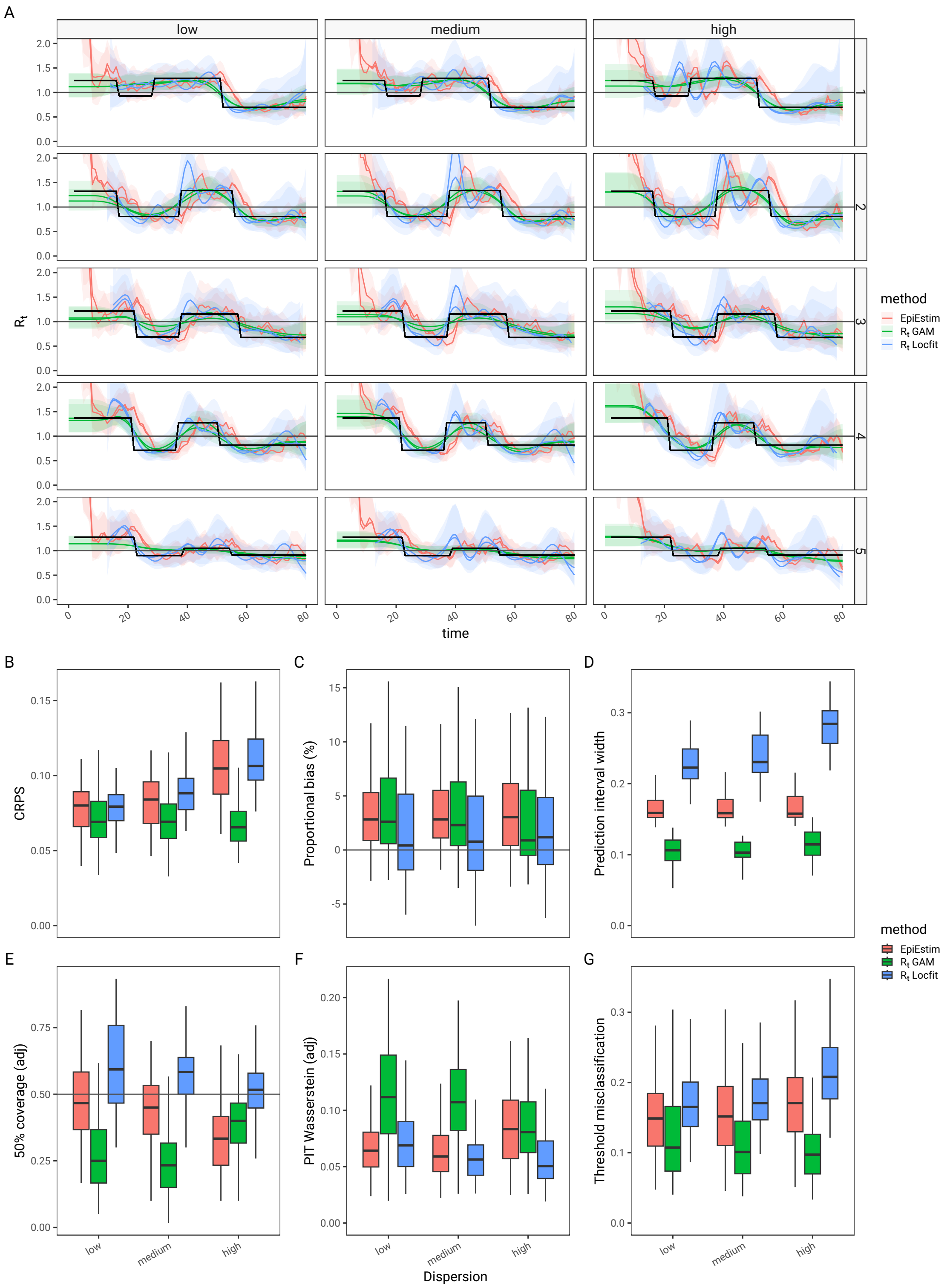


Figure 5: Sensitivity analysis of 'EpiEstim', and '$R_t$ from incidence' estimates derived from 7 day data windows, as opposed to 14, as described in the main paper, and are shown in Panel A for each of 5 scenarios (highest and lowest replicates only). The figure compares the continuous ranked probability score (CRPS) - lower is better, panel B; the average proportional bias, in panel C - zero is best; The 50% prediction interval width which measures estimator sharpness and lower is better, so long as the estimator is unbiased and well calibrated (panel E); the probability of 50% coverage (ideal value is 0.5, panel E), and adjusted probability integral transform (PIT) Wasserstein metric (lower is better, panel F); the probability of misclassifying at the threshold of $R_t = 1$ is shown in panel G (lower is better).

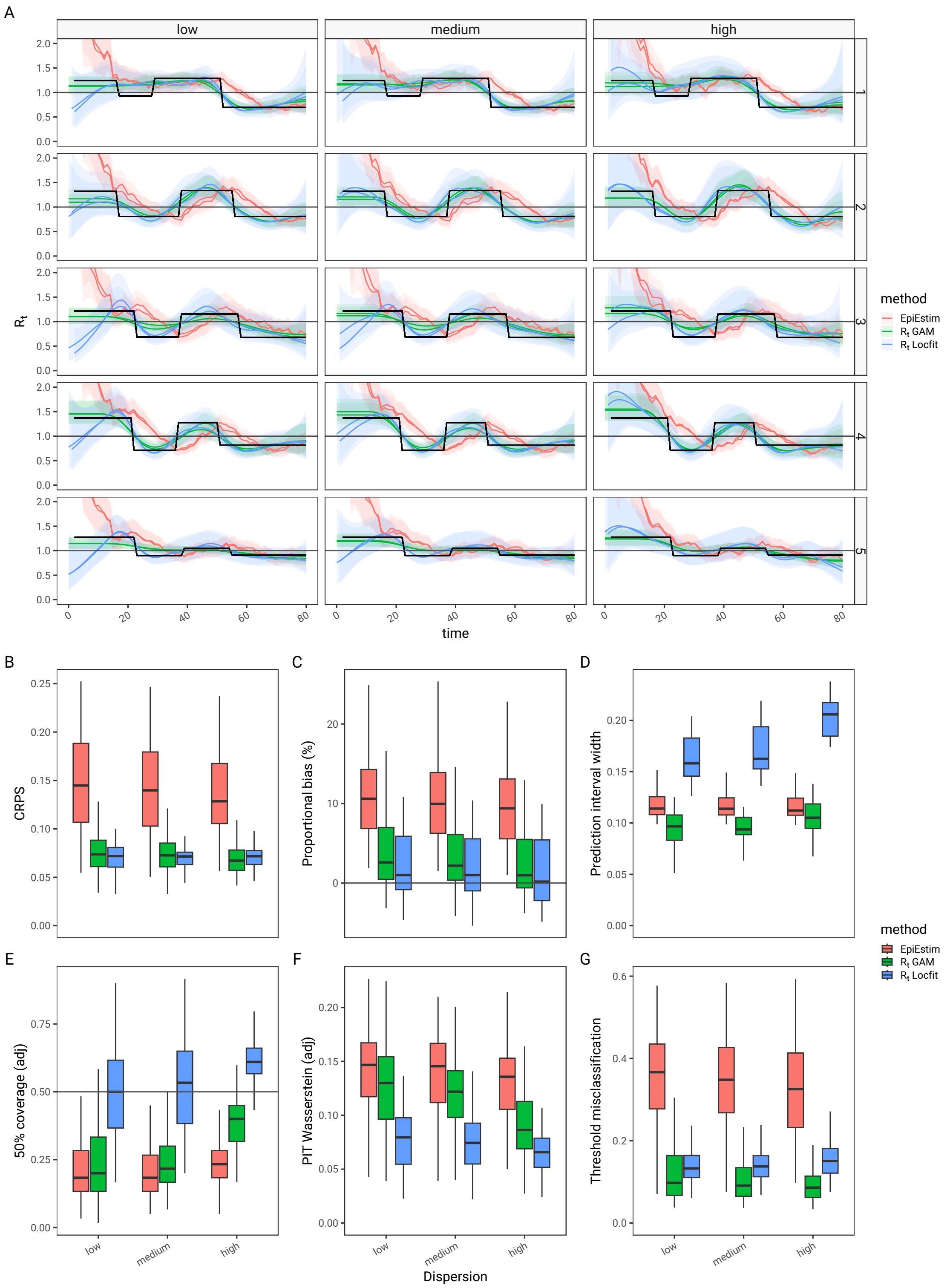


Figure 6: Sensitivity analysis of 'EpiEstim', and '$R_t$ from incidence' estimates when neither estimate is corrected for lag, are shown in Panel A for each of 5 scenarios (highest and lowest replicates only). The figure compares the continuous ranked probability score (CRPS) - lower is better, panel B; the average proportional bias, in panel C - zero is best; The 50% prediction interval width which measures estimator sharpness and lower is better, so long as the estimator is unbiased and well calibrated (panel E); the probability of 50% coverage (ideal value is 0.5, panel E), and adjusted probability integral transform (PIT) Wasserstein metric (lower is better, panel F); the probability of misclassifying at the threshold of $R_t = 1$ is shown in panel G (lower is better).

### References

[1] Katelyn M. Gostic, Lauren McGough, Edward B. Baskerville, Sam Abbott, Keya Joshi, Christine Tedijanto, Rebecca Kahn, Rene Niehus, James A. Hay, Pablo M. De Salazar, Joel Hellewell, Sophie Meakin, James D. Munday, Nikos I. Bosse, Katharine Sherrat, Robin N. Thompson, Laura F. White, Jana S. Huisman, Jérémie Scire, Sebastian Bonhoeffer, Tanja Stadler, Jacco Wallinga, Sebastian Funk, Marc Lipsitch, and Sarah Cobey. Practical considerations for measuring the effective reproductive number, Rt. 16(12):e1008409. URL: https://journals.plos.org/ploscompbiol/article?id=10.1371/journal.pcbi.1008409, doi:10.1371/journal.pcbi.1008409.

[2] R. N. Thompson, J. E. Stockwin, R. D. van Gaalen, J. A. Polonsky, Z. N. Kamvar, P. A. Demarsh, E. Dahlqwist, S. Li, E. Miguel, T. Jombart, J. Lessler, S. Cauchemez, and A. Cori. Improved inference of time-varying reproduction numbers during infectious disease outbreaks. 29:100356. URL: https://www.sciencedirect.com/science/article/pii/S1755436519300350, doi:10.1016/j.epidem.2019.100356.

[3] Kris V. Parag. Improved estimation of time-varying reproduction numbers at low case incidence and between epidemic waves. 17(9):e1009347. URL: https://journals.plos.org/ploscompbiol/article?id=10.1371/journal.pcbi.1009347, doi:10.1371/journal.pcbi.1009347.
