## Supplementary material for "Instantaneous Reproduction Number Estimation From Modelled Incidence": S2 Appendix

#### 1 Background

This appendix describes the use of metrics to characterise probabilistic estimators. We define an estimator as an algorithm that uses data to make probabilistic predictions of real world variables. Each prediction is expressed as a parameterised probability distribution or as a set of quantiles. For the purpose of this discussion we will assume we know the true value of these variables, either from simulation, or from a held out or delayed set of observations. We wish to measure the quality of the probabilistic estimator, through the set of true values and their associated predictions.

A probabilistic estimator has three aspects which determine its utility. Firstly it can have a bias, where the value of the predictions may be systematically higher or lower than the true values. Secondly it will have an sharpness which describes on average how precise each individual prediction will be. Thirdly a probabilistic estimator will have a probabilistic calibration, which describes whether, on average, the estimate uncertainty is representative of the dispersion of the estimates. A poorly calibrated estimator may produce estimates that are imprecise (too conservative) or over-precise (over-confident), leading in the first instance to predictions that are always right but uninformative, and in the second instance to predictions that may be misleading.

In the special case of time series estimators, there may be a systematic mismatch between the timing of prediction and true value giving a lag error. These are relatively easy to characterise with a cross-correlation function and correct by shifting the true values or the estimates in time and not considered further in this appendix. There may also be temporal patterns to errors which are out of scope of this paper.

The bias of an estimator is simply the average of the difference between central values of each prediction and true value. In our framework we use the median as the central value as the most commonly reported statistic. The sharpness of an individual probabilistic prediction is described by the average of the variance of the prediction, or by the width of a prediction interval (e.g. an interval of 50%, which is the inter-quartile range). The average sharpness is a properly of the estimator and describes how confident the estimator is. The calibration of a prediction can be thought of as a proxy for the spread of the error function of the estimator (see Fig 1), and measures how appropriate the estimator's confidence is. There is a trade off between bias, sharpness and calibration, and combined metrics aim to measure overall performance combining all three aspects.

The purpose of this appendix is to describe the metrics used in the main paper to characterise

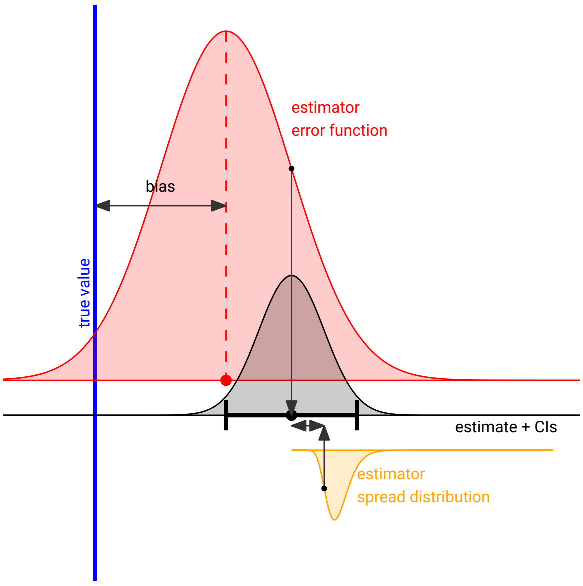


Figure 1: An estimator generates probabilistic predictions for any given true values (blue line), based on data (not shown). The estimator has an inherent propensity for error (red curve). For any given true value the central value of a probabilistic prediction (black curve) may not match the true value, and if this is consistently over many predictions, the estimator may be regarded as biased. The estimator also generates a measure of uncertainty of the probabilistic prediction (black bars). Over many predictions the distribution of this uncertainty is a measure of estimator sharpness (yellow curve). The sharpness ideally matches the spread of the estimator's error function. If, as is shown here, the estimators sharpness is narrower than the error function spread then any single prediction is likely to be over-confident. This is the result of a mis-calibrated and over confident estimator. The ratio of the error function standard deviation and the median of the estimator spread distribution (sharpness) is the true value of the calibration.

the quality of estimators in terms of bias, sharpness and calibration. For calibration we introduce and characterise a novel measure that is not affected by estimator bias and sharpness, which is useful for comparing the calibration of different estimators.

#### 2 Methods

In this paper we describe a set of true values of a measure with the notation $X_i$ where $i \in 1 \ldots n$. The probabilistic prediction of this true value is a function which we describe with the notation $F_i(x) = Pr(X_i \leq x)$. We describe the quantiles of this distribution using the notation $x_{i,0.5}$ where $F_i(x_{i,0.5}) = 0.5$ and so on. For simplicity sake we define $X_i$ to be in ascending order $(X_i \leq X_{i+1})$.

A well described combined metric for measuring the quality of a probabilistic estimator that encompasses bias, sharpness and calibration is the average of the Continuous Rank Probability Score (CRPS) of each prediction [1, 2, 3, 4, 5]. This is defined as follows where $H$ is the Heaviside step function, and smaller values imply better predictions:

$$
\begin{aligned}
CRPS_i &= \int_{\mathbb{R}} (F_i(x) - H(x < X_i))^2 dx \\
\overline{CRPS} &= \frac{1}{n} \sum_{n} CRPS_i
\end{aligned} \tag{1}
$$

The average value of every prediction's CRPS is a measure of the estimator itself and this is a proper scoring rule, in that it is minimised by a sharp, well calibrated unbiased estimator. It is, however, useful when comparing estimators to characterise bias, sharpness and calibration individually.

For a linear variable the bias of an estimator is simply the average difference between a set of true values and the central quantile of the predictions. For logarithmic variables a better representation is the average proportional bias. If we specify a link function $(g)$ that relates the variable of interest to a linear scale we can generalise the estimator bias $(\overline{\mathcal{E}_X})$ as follows, and with this value defined we can define an adjusted true value $X_{i,adj}$.

$$\begin{aligned} \overline{\mathcal{E}_X} &= \frac{1}{n}\sum_n g(X_i) - g(x_{i,0.5}) \\ X_{i,adj} &= g'(g(X_i) - \mathcal{E}_X) \end{aligned} \tag{2}$$

In the case of the reproduction number described in the main paper the link function is $g(R_t) = log(R_t)$ and the resulting bias is multiplicative and represented as the percentage $e^{\mathcal{E}_{R_t}} - 1$.

When individual predictions are available as posterior continuous probability distributions, they can be assessed using probability integral transform (PIT) values $u_i$ [5, 6] which are the estimated cumulative distribution function (CDF, $F_i(x)$) evaluated at the true values of $X_i$. PIT values (also sometimes described as universal residuals [7]) $u_i$. The average universal residual, rescaled to range from -1 to 1, $\overline{\mathcal{E}_u}$ gives an indication of estimator bias that is independent of the distributional form of the estimator.

$$\begin{aligned} u_i &= F_i(X_i) \\ \overline{\mathcal{E}_u} &= \frac{1}{n}\sum (2u_i - 1) \end{aligned} \tag{3}$$

The sharpness of an probabilistic estimator can be measured as the average variance of predictions, or, in the main paper, as the average width of predictions at a given level of probability (usually 50% - $\overline{\omega_{0.5}}$). This is independent of the true value.

$$\overline{\omega_{0.5}} = \frac{1}{n}\sum (x_{i,0.75} - x_{i,0.25}) \tag{4}$$

For a single prediction we can assign a logical value to the condition that the true value is between the 0.25 and 0.75 quantiles. For a set of predictions produced by a single estimator the average gives us the probability of 50% coverage as a metric for the the calibration of the estimator $(\mathcal{P}_{X,0.5})$. In a well calibrated unbiased estimator the probability that the true value is within the quartiles should be 0.5, whereas over confident estimators will score lower, and excessively conservative higher. Bias affects the 50% coverage probability, as a shift in the quartiles reduces the chance that the true value lies within the range.

$$\mathcal{P}_{X,0.5} = \frac{1}{n}\sum_n 1(0.25 \leq F_i(X_i) \leq 0.75) \tag{5}$$

We defined the probability integral transform (PIT) for a single prediction above. For many predictions if the estimator is well calibrated the PIT value should be approximately uniformly distributed on [0,1]. It is common practice to visually compare PIT histograms and look for flatness. If the PIT value histogram is U-shaped, and there is more mass at the edges, it represents an over confident prediction, or if dome shaped with more mass in the centre, it represents an excessively uncertain prediction. However if bias is also present the PIT histogram is then skewed and difficult to interpret and compare [8, 1].

To address this and to quantify the difference between the distribution of the PIT values and the uniform, we initially adjust the estimates for bias and then calculate an adjusted PIT value for each predictions $u_{i,adj}$. We summarise these using a Wasserstein distance, (also known as earth movers cost) [9] between the PIT values and the ideal uniform distribution. This adjusted PIT Wasserstein metric ($D^*_{pit}$) is strictly positive, will take a maximum value of 0.5 if all the mass of the PIT distribution is centred at any one point, and a minimum value of zero if the estimator is perfectly calibrated.

$$\begin{aligned} rank_i &= \frac{i - 0.5}{n} \\ u_{i,adj} &= F_i(X_{i,adj}) \\ D^*_{pit} &= \frac{1}{n}\sum_{i=1}^{n} |rank_i - u_{i,adj}| \end{aligned} \tag{6}$$

We can calculate another PIT centrality measure ($U^*_{pit}$) as Wasserstein distance from the PIT distribution to the uniform distribution with positive cost if moving away from the centre, and negative cost if moving towards the centre. This directed PIT Wasserstein metric will take values between -0.25 if all the mass is at the edges representing over-confidence in predictions, to 0.25 if all the mass is at the centre, representing excessive uncertainty in predictions. An ideal estimator would have zero for this metric:

$$U^*_{pit} = \frac{1}{n}\sum_{i=1}^{n} (rank_i - u_{i,adj}) \times \operatorname{sgn}(0.5 - rank_i) \tag{7}$$

Whilst these generic metrics are informative, the main value of an $R_t$ estimate is to answer the question “is the epidemic increasing?” and a metric for $R_t$ comparison which describes how accurately, and with what confidence, the estimator predicts when the $R_t$ value is greater or less than 1, is useful. In some situations it may be preferable to have an over sharp, poorly calibrated estimate if it is better at predicting the growth phases of the epidemic. We consider the probability of an erroneous conclusion at a threshold value $T$ for an individual prediction, compared to the true value ($\mathcal{P}_{i,err}$). If we combine these for an estimator, using a weighted average, where the weight is the distance of the true value from the threshold on the link function scale then we can define a threshold misclassification probability ($\mathcal{P}_{err}$) as the probability the estimator will produce an prediction that can be misinterpreted against the threshold criteria, which in the case of $R_t$ is above or below 1:

$$\begin{aligned} \mathcal{P}_{i,err} &= |F_i(T) - 1(X_i \leq T)| \\ \mathcal{P}_{err} &= \frac{\sum \mathcal{P}_{i,err} \times |g(X_i) - g(T)|}{\sum |g(X_i) - g(T)|} \end{aligned} \tag{8}$$

The threshold misclassification probability is relevant to variables such as $R_t$ and growth rate, but less so for incidence or prevalence, which do not have a natural threshold. It is a probability, and lower values are better for decision making. It will penalise situations where there is overt growth or decline but the prediction is either incorrect or is correct but excessively uncertain.

#### 2.1 Validation

To test metrics related to calibration, we have constructed a simple simulation, in which true values ($X_i$) are random samples from a standard normal distribution. A hypothetical estimator creates normally distributed probabilistic predictions of the true values, defined by the CDF $F_i$, offset from the true value by a random sample $\mathcal{E}$ from a normally distributed error function with mean $\mu_{\mathcal{E}}$ and standard deviation of $\sigma_{\mathcal{E}}$. The estimate uncertainty, as defined by $\sigma_i$ is a log-normally distributed sharpness parameterised by median $\sigma_{\mathcal{S}}$ and coefficient of variation $\kappa_{\mathcal{S}}$.

$$\begin{aligned} X &\sim N(0,1) \\ \mathcal{E} &\sim N(\mu_{\mathcal{E}}, \sigma_{\mathcal{E}}) \\ \log(\sigma_i) &\sim N\left(\log(\sigma_{\mathcal{S}}), \sqrt{\log(\kappa_{\mathcal{S}}^2 + 1)}\right) \\ F_i(x) &= \Phi\left(\frac{x - X_i - \mathcal{E}_i}{\sigma_i}\right) \end{aligned} \tag{9}$$

Where $\Phi$ is the standard normal distribution cumulative density function. With this arrangement the estimator bias is defined as $\mu_{\mathcal{E}}$, sharpness as $\sigma_{\mathcal{S}}$ and calibration is $\frac{\sigma_{\mathcal{S}}}{\sigma_{\mathcal{E}}}$. $\kappa_{\mathcal{S}}$ is a measure of the intrinsic variability of estimate confidence, and is an index of the variability of the confidence of predictions produced by a single estimator.

We ran this simulation 1000 times with random configurations, and calculated the metrics described above, based on 10000 simulated true value and probabilistic prediction pairs, for each of the 1000 random simulations. The summary estimator metrics were calculated without resampling.

$$\begin{aligned} \mu_{\mathcal{E}} &\sim Uniform(-1,1) \\ log(\sigma_{\mathcal{E}}) &\sim Uniform(-log(2), log(2)) \\ log(\sigma_{\mathcal{S}}) &\sim Uniform(-log(2), log(2)) \\ \kappa_{\mathcal{S}} &\sim Uniform(0,2) \end{aligned} \tag{10}$$

#### 2.2 Results and Discussion

The simulation was run for a curated set of configuration parameters shown in Table 1 and the resulting PIT histograms plotted in Fig 2. These demonstrate the expected PIT histogram shapes in the presence of bias and calibration errors, showing U shaped histograms for over-confident estimators, and dome shapes for excessively conservative ones. Skew in the PIT histograms is exhibited by configurations with positive or negative bias. In each plot the associated values of the adjusted PIT-Wasserstein (adj PIT-W), and directed PIT-Wasserstein (dir PIT-W) metrics, along with the mean CRPS for each of the represented estimators is given. The adjusted PIT-W metrics are invariant in the presence of bias, and appear correlated to calibration.

In Fig 3 we see the results of the 1000 random simulations, and the values of 4 estimator metrics are plotted as they relate to the theoretical value of the calibration on the x-axis ($\frac{\sigma_{\mathcal{S}}}{\sigma_{\mathcal{E}}}$), based on the simulation configurations defined in (10). The points are coloured according to

Table 1: Curated parameters for 9 simulated estimators with fixed sharpness and variable calibration and bias.

| biased | confidence | bias ($\mu_e$) | sharpness ($\sigma_e$) | calibration ($\sigma_e/\sigma_s$) | kappa ($\kappa$) |
|---|---|---|---|---|---|
| negatively | conservative | -0.50 | 1.00 | 1.50 | 0 |
| | appropriate | -0.50 | 1.00 | 1.00 | 0 |
| | overconfident | -0.50 | 1.00 | 0.67 | 0 |
| unbiased | conservative | 0.00 | 1.00 | 1.50 | 0 |
| | appropriate | 0.00 | 1.00 | 1.00 | 0 |
| | overconfident | 0.00 | 1.00 | 0.67 | 0 |
| positively | conservative | 0.50 | 1.00 | 1.50 | 0 |
| | appropriate | 0.50 | 1.00 | 1.00 | 0 |
| | overconfident | 0.50 | 1.00 | 0.67 | 0 |

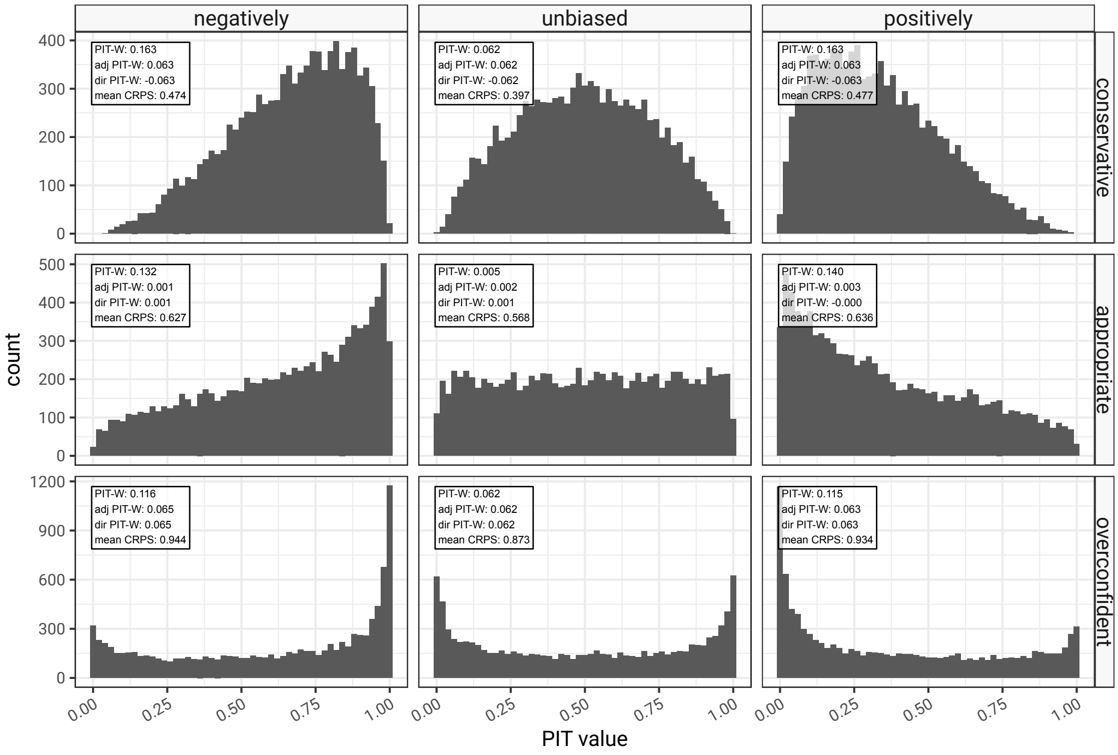


Figure 2: PIT histograms from 10000 simulated predictions from 9 simulated estimators with curated parameters, as given in Table 1. Typical U and dome shaped patterns are seen as a result of mis-calibration, and skew as a result of estimator bias

the variation in confidence intervals represented by the value of $\kappa_S$. In panel A the CRPS is correlated with calibration, but does not exhibit a minimum at a well calibrated, and tends to favour conservative estimates. The CRPS is composite metric and affected by bias and sharpness, as well as by calibration, which can compensate for bias when the estimator is conservative. The 50% coverage probability in Fig 3 panel B is used as a metric for calibration, and demonstrates a better relationship with calibration, but it too is affected by bias, as shifted estimates are less likely to contain the true value, unless they are also excessively conservative. In the presence of minor bias coverage probability is likely to give a good indication of the direction and to a lesser extent the scale of mis-calibration. In Fig 3 panel C we introduce the adjusted PIT-Wasserstein metric which has a clear relationship with calibration that is not affected by bias or sharpness. It appears to be a proper scoring rule for calibration and is symmetric on the logarithm of calibration. This means that an estimator that produces estimates that have confidence intervals twice as large as they should be will score the same as one whose confidence intervals are half as wide as they should be. This is a useful property for comparisons between estimators. The directed PIT-Wasserstein metric in Fig 3 panel D is similarly unaffected by bias and sharpness, and similar to the undirected version but gives us a clear direction of mis-calibration similar to the 50% coverage probability.

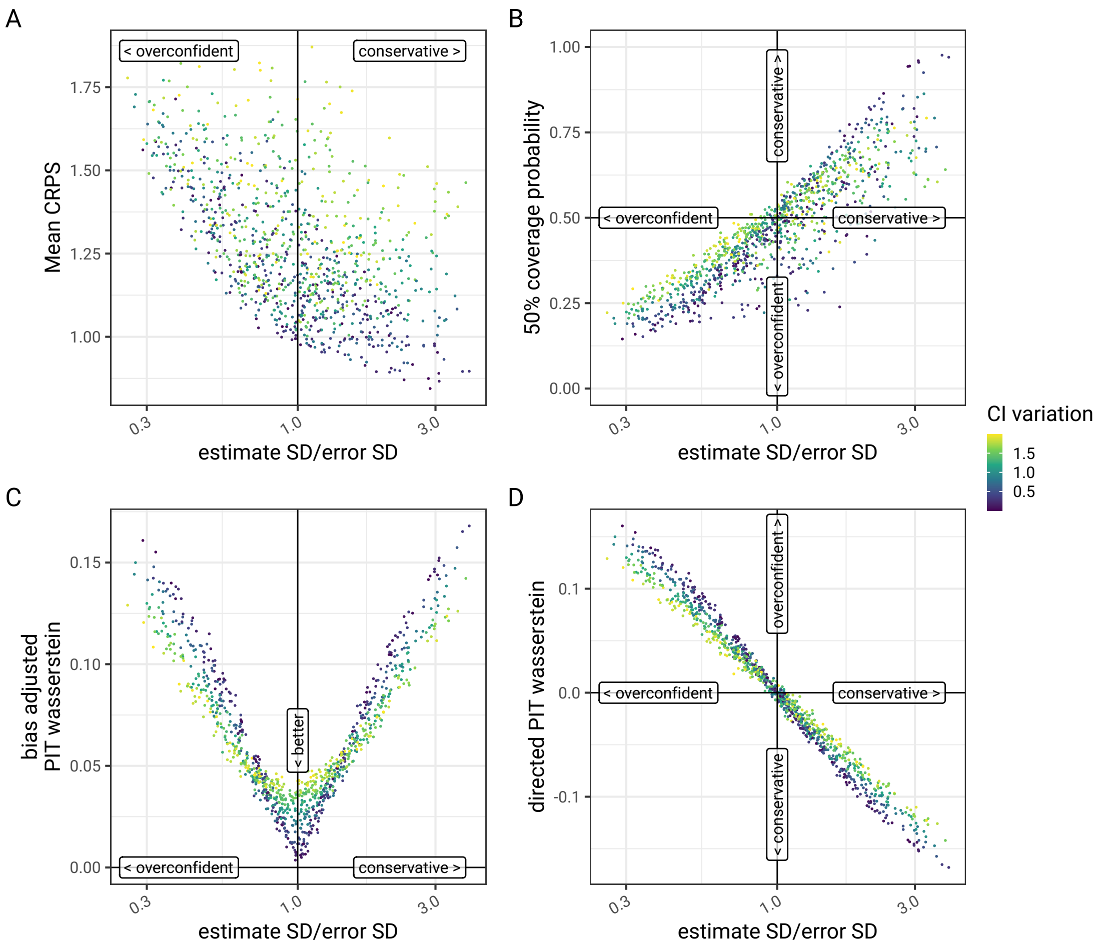


Figure 3: Comparison of four estimator metrics for 1000 random simulated estimators, consisting of 10000 synthetic predications, as it relates to the theoretical calibration of the estimator on the X-axes. Panel A shows the mean CRPS, panel B the 50% coverage probability. Panels C and D show adjusted and directed PIT-Wasserstein metrics, as described earlier in this paper, which are designed to give a measure of calibration that is independent of estimator bias and sharpness.

Fig 4 demonstrates the effect of removing bias on the mean CRPS and 50% coverage probability and serves to underline what was said previously about the fact these metrics are not independent representations of calibration. However at least in low bias situations the 50% coverage probability is a serviceable option, and well understood.

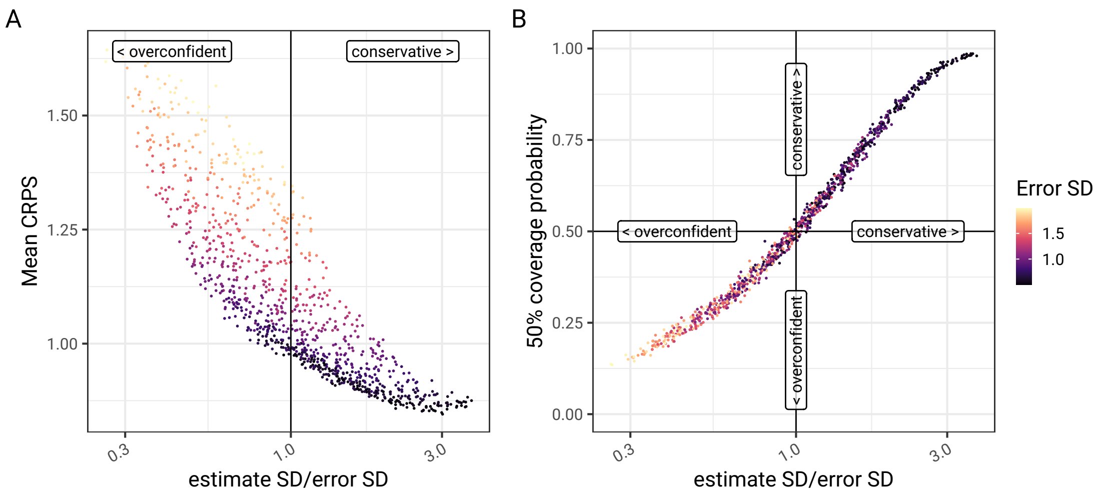


Figure 4: A repeated comparison of traditional probabilistic estimator metrics, mean CRPS (Panel A) and 50% coverage probability (Panel B) in the restricted scenario where simulated estimators are unbiased. In the absence of bias, the 50% coverage probability is a good representation of calibration. The mean CRPS also is affected by sharpness so has unresolved interactions.

### 3 Conclusion

In this paper we introduce a novel metric of probabilistic estimator calibration, the adjusted PIT-Wasserstein distance. We show that this works well to specifically identify issues with calibration independent of bias and sharpness. Further work is required to characterise the behaviour of this in small samples, and to identify thresholds for significance.

We describe the selection of metrics that we use to compare probabilistic estimators of $R_t$ in the main paper, and provide a formal definition. These are summarised as follows.

- Overall:
  - the average continuous rank probability score (CRPS). Lower values imply better predictions.
- Bias:
  - the exponent of the average error on a logarithmic scale. Like $R_t$ this is a multiplicative measure and values closer to 1 are better.
  - the average of the universal residuals. Values closer to zero are better.
- Sharpness:
  - the 50% prediction interval width. Lower values are better but only if the estimate is unbiased and correctly calibrated.
- Calibration:
  - the 50% coverage probability. Ideally this is around 0.5, with lower values representing over-confidence in the estimator and larger values representing an excessively conservative estimator. Bias can affect this metric, and an alternative is the directed PIT-Wasserstein distance.

- the adjusted PIT-Wasserstein distance. In general lower values are better, but the trade off between sharpness and calibration has to be considered.

- Utility:
  - the threshold misclassification probability. Lower values are better. High values penalise the proportion of an estimate that is wrongly on the other side of the threshold, scaled by the distance of the true value from the threshold. This will penalise estimators that provide evidence for the wrong outcome, and hence favours confident correct estimators even if they are too sharp.

[2] Nikos I. Bosse, Hugo Gruson, Anne Cori, Edwin van Leeuwen, Sebastian Funk, and Sam Abbott. Evaluating Forecasts with scoringutils in R. URL: `http://arxiv.org/abs/2205.07090`, `arXiv:2205.07090`, `doi:10.48550/arXiv.2205.07090`.

[3] Nikos I. Bosse, Sam Abbott, Anne Cori, Edwin van Leeuwen, Johannes Bracher, and Sebastian Funk. Scoring epidemiological forecasts on transformed scales. 19(8):e1011393. URL: `https://journals.plos.org/ploscompbiol/article?id=10.1371/journal.pcbi.1011393`, `doi:10.1371/journal.pcbi.1011393`.

[4] J. Bröcker. On reliability analysis of multi-categorical forecasts. 15(4):661–673. URL: `https://npg.copernicus.org/articles/15/661/2008/`, `doi:10.5194/npg-15-661-2008`.

[5] Tilmann Gneiting and Adrian E Raftery. Strictly Proper Scoring Rules, Prediction, and Estimation. 102(477):359–378. `doi:10.1198/016214506000001437`.

[6] F. N. David and N. L. Johnson. The Probability Integral Transformation When Parameters are Estimated from the Sample. 35(1/2):182–190. URL: `https://www.jstor.org/stable/2332638`, `arXiv:2332638`, `doi:10.2307/2332638`.

[7] A. E. Brockwell. Universal residuals: A multivariate transformation. 77(14):1473–1478. URL: `https://www.sciencedirect.com/science/article/pii/S0167715207000715`, `doi:10.1016/j.spl.2007.02.008`.

[8] Thomas M. Hamill. Interpretation of Rank Histograms for Verifying Ensemble Forecasts. 129(3):550–560. URL: `https://journals.ametsoc.org/view/journals/mwre/129/3/1520-0493_2001_129_0550_iorhfv_2.0.co_2.xml`, `doi:10.1175/1520-0493(2001)129<0550:IORHFV>2.0.CO;2`.

[9] Victor M. Panaretos and Yoav Zemel. Statistical Aspects of Wasserstein Distances. 6:405–431. URL: `https://www.annualreviews.org/content/journals/10.1146/annurev-statistics-030718-104938`, `doi:10.1146/annurev-statistics-030718-104938`.
