## Supplementary material for "Instantaneous Reproduction Number Estimation From Modelled Incidence": S3 Appendix

### Supplementary 3: Bias in $R_t$ Estimation Under Independence and Heuristics for Risk Assessment

#### 1 Background

When the full posterior variance-covariance (VCOV) matrix of log-incidence estimates is unavailable, it is common to assume that uncertainty in $\lambda_t$ is independent over time. This greatly simplifies computation, and as such it could be a pragmatic assumption to make even if a VCOV matrix is available, but it can introduce bias in $R_t$ due to misestimation of the denominator's variance and its covariance with the numerator.

This note quantifies the bias in $R_t$ under the independence assumption, and derives heuristics for detecting high-risk scenarios.

In the situation that a VCOV matrix is not available from the esitmation framework we describe methods to approximate the VCOV matrix from residuals.

#### 2 Bias from assuming independence

To recap the method described in the main paper, the renewal equation gives:

$$R_t \approx \frac{\lambda_t}{\sum_{\tau=1}^{k} \omega_\tau \lambda_{t-\tau}} = \frac{\lambda_t}{S_t} \tag{1}$$

where $S_t$ is the expected infectious pressure from past cases. Using the Lo et al. [1] approximation, $S_t \approx \text{Lognormal}(\mu_Z, \sigma_Z)$, with:

$$\begin{aligned} \mu_Z &= \log S_+ - \frac{1}{2}\sigma_Z^2 \\ m_\tau &= E[\omega_\tau \lambda_{t-\tau}] = e^{\left(\mu_{t-\tau} + \log \omega_\tau + \frac{1}{2}\sigma_{t-\tau}^2\right)} \\ S_+ &= \sum_{\tau=1}^{k} m_\tau \end{aligned} \tag{2}$$

An approximation of the true variance of $\log S_t$ is:

$$\sigma_Z^2 = \frac{1}{S_+^2} \sum_{i,j=1}^{k} m_i m_j \mathbb{V}_{ij} \quad \text{where} \quad \mathbb{V}_{ij} = \text{Cov}(\log \lambda_{t-i}, \log \lambda_{t-j}) \tag{3}$$

This leads to a formulation of the distributional form of $R_t$ as:

$$\begin{aligned}
R_t &\sim \frac{Lognormal(\mu_t, \sigma_t)}{Lognormal(\mu_Z, \sigma_Z)} = Lognormal(\mu_{R_t}, \sigma_{R_t}) \\
\mu_{R_t} &= \mu_t - \mu_Z = \mu_t - \log S_+ + \frac{1}{2}\sigma_Z^2 \\
\sigma_{R_t} &= \sqrt{\sigma_t^2 + \sigma_Z^2 - 2\mathbb{V}_{0Z}} \\
\mathbb{V}_{0Z} &= \text{Cov}(\log \lambda_0, \log S_t) \approx \frac{1}{S_+}\sum_{\tau=1}^{k} m_\tau \mathbb{V}_{0\tau}
\end{aligned} \tag{4}$$

Under the assumption of independence we can calculate a naive estimate of $R_t$ where $\mathbb{V}_{ij} = \delta_{ij}\sigma_{t-i}\sigma_{t-j}$ and $\mathbb{V}_{0Z} = 0$. From the definition of $\mu_{R_t}$ we can also see it will be potentially biased. We can quantify this bias by looking at the ratio of median of true and 'naive' estimates made under the assumption of independence, $B_{median}$, as follows:

$$\begin{aligned}
B_{median} &= \frac{\text{Median}[R_{t,true}]}{\text{Median}[R_{t,naive}]} = \frac{e^{(\mu_{R_{t,true}})}}{e^{(\mu_{R_{t,naive}})}} \\
\beta_{median} = \log B_{median} &= \mu_t - \log S_+ + \frac{1}{2}\sigma_{Z,true}^2 - (\mu_t - \log S_+ + \frac{1}{2}\sigma_{Z,naive}^2) \\
\beta_{median} &= \frac{1}{2}(\sigma_{Z,true}^2 - \sigma_{Z,naive}^2) = \frac{1}{2}\Delta\sigma_Z^2 \\
\sigma_{Z,naive}^2 &= \frac{1}{S_+^2}\sum_{\tau=1}^{k} m_\tau^2 \sigma_{t-\tau}^2 \\
\beta_{median} = \Delta\sigma_Z^2 &= \frac{\sum_{i\neq j} m_i m_j \mathbb{V}_{ij}}{S_+^2}
\end{aligned} \tag{5}$$

$\sigma_{Z,true}^2$ includes off-diagonal terms compared to $\sigma_{Z,naive}^2$ and this leads to a systematic bias in the median $R_t$ estimates by a factor of $[\exp(\Delta\sigma_Z^2) - 1]$. This is a strictly positive term, implying that assuming independence causes median estimates of $R_t$ to be underestimated. It is not shown here but mean estimates of $R_t$ will be less biased and could potentially be overestimated in some unusual situations.

If we do not have information on the correlation structure we can estimate the risk and magnitude of this bias using the weighted average variance ($V_+$), which is the effective uncertainty in the denominator components, and the maximum possible off-diagonal contribution to $\sigma_Z^2$, achieved under perfect correlation ($c_{\max}$).

$$\begin{aligned}
V_+ &= \sum_{\tau=1}^{k} \frac{m_\tau}{S_+}\sigma_{t-\tau}^2 \\
c_{\max} &= 1 - \sum_{\tau=1}^{k} \frac{m_\tau}{S_+}^2
\end{aligned} \tag{6}$$

Then the maximum possible bias in $R_t$ is:

$$
\begin{aligned}
\max(\beta_{median}) &\approx \frac{1}{2}c_{\max}V_{+} \\
\Delta R_t &= \text{Median}[R_{t,true}] - \text{Median}[R_{t,naive}] \\
\max(\Delta R_t) &= \text{Median}[R_{t,naive}](e^{\frac{1}{2}c_{\max}V_{+}} - 1)
\end{aligned}
\tag{7}
$$

We know this worst case scenario is moderated by the weight of the off diagonal covariance terms weighted by the infectivity profile, and that an estimate for the total weight of the variance-covariance is given by $V_{+}$. With this we can estimate the fractional contribution of the unaccounted for covariance:

$$
\text{Risk of bias} = \frac{V_{+}}{\sigma^2_{Z,\text{naive}}} \tag{8}
$$

With these values we can identify specific situations where the bias is unacceptable, and in particular the relative error in $R_t$ estimates. Typically a maximum underestimate of 5% will be tolerable given other uncertainties.

$$
\begin{aligned}
0.05 &\geq e^{\frac{1}{2}c_{\max}V_{+}} - 1 \\
0.1 &\gtrsim c_{\max}V_{+} \gtrsim \overline{\sigma_t}^2
\end{aligned}
\tag{9}
$$

In this expression $c_{\max}$ is a unitless number between 0 and 1, and $V_{+}$ is a weighted average of the square of the $\sigma$ parameters of our log normal incidence estimates. If the estimates for $\sigma_t$ are stable we can ignore the weighting and be confident that if the average $\overline{\sigma_t}$ is less than 0.3 then the magnitude of bias is acceptable. Secondly the off diagonal contribution $c_{\max}$ may be small, potentially due to a peaked infectivity profile, or stable incidence. Even in the situation where the maximum possible bias is large there may be mitigation due to the structure of the covariance matrix.

In an integrated framework we can assess the risk of bias for every derived $R_t$ estimate when assuming independence and flag them as high risk if both the magnitude and the risk is high. and specifically if $\max(\Delta R_t) > 0.05$ and $\frac{V_{+}}{\sigma^2_{Z,\text{naive}}} > 5$.

### 3 Estimating VCOV from Residuals

If the VCOV matrix is not available from the incidence model, but raw data $I_t$ and fitted $\hat{\lambda}_t$ are, we can estimate a synthetic VCOV using Pearson residuals $\nabla_t$ and compute the sample autocorrelation $\hat{\rho}_\tau$, and fit an exponential decal model by nonlinear least squares or method of moments:

$$
\begin{aligned}
\nabla_t &= \frac{I_t - \hat{\lambda}_t}{\sqrt{\hat{\lambda}_t}} \\
\hat{\rho}_\tau &= \text{Corr}(\nabla, \nabla_{t-\tau}) \\
\rho_\tau &\sim \mathit{Exponential}(-\alpha\tau) \quad \text{for } \tau = 1, \ldots, k
\end{aligned}
\tag{10}
$$

A synthetic VCOV matrix can then be reconstructed which captures the smoothness-induced correlation typical in GAMs and smoothing models. This estimates the data-level correlation, not full posterior uncertainty, but is sufficient for bias correction.

$$\mathbb{V}_{ij} = e^{-\alpha|j-i|} \cdot \sigma_{t-i}\sigma_{t-j} \tag{11}$$

### 4 Conclusion

While the independence assumption can introduce downward bias in $R_t$, the magnitude is often small in practice. Because the bias is related to the uncertainty of the denominator, any bias will be also matched by an increase in the uncertainty of the $R_t$ estimate, which will tend to compensate in interpretation. In implementation we can save computational time by supporting the assumption of independence and detecting if the risk of bias is unacceptable. This also supports statistical frameworks that do not produce estimate VCOV matrices. In this latter case if the origial data is available we can further support these by providing the option to use a synthetic VCOV matrix.

### References


[1] Chi-Fai Lo. WKB Approximation for the Sum of Two Correlated Lognormal Random Variables. 7(129):6355–6367. URL: https://papers.ssrn.com/abstract=2220803, doi:10.2139/ssrn.2220803.
